# Longitudinal changes in Alzheimer’s-related plasma biomarkers and brain amyloid

**DOI:** 10.1101/2023.01.12.23284439

**Authors:** Murat Bilgel, Yang An, Keenan A. Walker, Abhay R. Moghekar, Nicholas J. Ashton, Przemysław R. Kac, Thomas K. Karikari, Kaj Blennow, Henrik Zetterberg, Bruno M. Jedynak, Madhav Thambisetty, Luigi Ferrucci, Susan M. Resnick

## Abstract

**Introduction:** Understanding longitudinal plasma biomarker trajectories relative to brain amyloid changes can help devise Alzheimer’s progression assessment strategies.

**Methods:** We examined the temporal order of changes in plasma amyloid-β ratio (Aβ_42_/Aβ_40_), glial fibrillary acidic protein (GFAP), neurofilament light chain (NfL), and phosphorylated tau ratios (p-tau181/Aβ_42_, p-tau231/Aβ_42_) relative to ^11^C-Pittsburgh compound B (PiB) positron emission tomography (PET) cortical amyloid burden (PiB−/+). Participants (n = 199) were cognitively normal at index visit with a median 6.1-year follow-up.

**Results:** PiB groups exhibited different rates of longitudinal change in Aβ_42_/Aβ_40_ (*β* = 5.41 × 10^-4^, SE = 1.95 × 10^-4^, *p* = 0.0073). Change in brain amyloid correlated with change in GFAP (*r* = 0.5, 95% CI = [0.26, 0.68]). Greatest relative decline in Aβ_42_/Aβ_40_ (-1%/year) preceded brain amyloid positivity by 41 years (95% CI = [32, 53]).

**Discussion:** Plasma Aβ_42_/Aβ_40_ may begin declining decades prior to brain amyloid accumulation, whereas p-tau ratios, GFAP, and NfL increase closer in time.

## 1 Background

Plasma biomarkers of Alzheimer’s disease (AD)-related pathology and neurodegeneration are proxies of these changes in the central nervous system. Their low cost and ease of collection make them good candidates for widespread clinical use for assessing AD-related changes.

Amyloid-β (Aβ) accumulation marks the beginning of preclinical Alzheimer’s among cognitively unimpaired individuals [1]. As highlighted in the research priorities outlined by Hansson et al. [2], it is important to understand longitudinal changes in plasma biomarkers relative to the onset of this hallmark neuropathology. A better understanding of longitudinal plasma biomarker trajectories can improve patient selection and monitoring in clinical trials, facilitating identification of individuals at high risk of developing neurodegenerative changes and cognitive impairment. Plasma biomarkers may be particularly useful in limiting the number of positron emission tomography (PET) scans conducted to determine participant eligibility for trials of anti-amyloid treatments [3–7].

Despite rapidly developing research on plasma biomarkers, studies investigating longitudinal change remain limited. Chatterjee et al. reported that plasma Aβ_42_/Aβ_40_, tau phosphorylated at threonine 181 (p-tau181), and glial fibrillary acidic protein (GFAP) change more rapidly among individuals with mild cognitive impairment (MCI) compared to cognitively normal individuals [8]. O’Connor et al. found that longitudinal trajectories of plasma neurofilament light chain (NfL) and p-tau181 among autosomal dominant AD mutation carriers started diverging from trajectories observed for non-carriers at about 16–17 years prior to estimated symptom onset [9]. Plasma Aβ_42_/Aβ_40_ [10] and p-tau181 [11] also exhibit changes prior to elevated brain amyloid levels, with plasma Aβ changing prior to p-tau181 [12]. In a cohort of individuals with and without cognitive impairment, Rauchmann et al. examined trajectories of plasma p-tau181 and NfL relative to cerebrospinal fluid (CSF) or imaging measure-based definitions of amyloid (A), tau (T), and neurodegeneration (N) status and found that relative to the A−TN− group, all other groups exhibited steeper longitudinal increases in NfL [13]. Further, recent cross-sectional and longitudinal studies have shown early changes of all plasma biomarkers but note that p-tau231 changes earliest in response to Aβ deposition [14–16]. These findings suggest that these plasma biomarkers may be dynamic in the preclinical phase of AD and even earlier. However, it remains unclear how closely longitudinal changes in plasma biomarkers mirror longitudinal changes in brain amyloid levels.

In this study, we focus on understanding the temporal order of changes in AD-related plasma biomarkers relative to brain amyloid levels as measured with ^11^C-Pittsburgh compound B (PiB) PET. The plasma measures we consider are Aβ_42_, Aβ_40_, GFAP, NfL, p-tau181, and p-tau231 concentrations as well as the ratios Aβ_42_/Aβ_40_, p-tau181/Aβ_42_, and p-tau231/Aβ_42_. In cross-sectional analyses, we first replicate previous findings regarding their accuracy in classifying amyloid PET status. We then use longitudinal data to quantify their longitudinal intraclass correlation coefficients, estimate their trajectories as a function of brain amyloid status, investigate the associations among longitudinal rates of change in plasma and brain amyloid measures, and finally, examine the temporal order of changes in plasma measures relative to elevation in cerebral fibrillar amyloid burden.

## 2 Methods

### 2.1 Participants

Our sample consisted of 199 initially cognitively normal Baltimore Longitudinal Study of Aging (BLSA) participants with both amyloid PET and plasma biomarkers. 176 participants had at least two visits with both amyloid PET and plasma biomarkers. 21% of participants developed MCI or dementia over the course of the study. Measurements at the index visit, defined as the earliest cognitively normal visit with a full set of plasma biomarkers, were used for cross-sectional analyses. All plasma biomarker measurements for these participants were used in longitudinal analyses, allowing for inclusion of visits where a subset of plasma biomarkers was missing (because measurement was not performed or did not meet quality control).

Research protocols were conducted in accordance with United States federal policy for the protection of human research subjects contained in Title 45 Part 46 of the Code of Federal Regulations, approved by local institutional review boards, and all participants gave written informed consent at each visit.

### 2.2 Cognitive assessment

Cognitively normal status was based on either (i) ≤ 3 errors on the Blessed Information-Memory-Concentration Test [17] and a Clinical Dementia Rating (CDR) [18] of zero, or (ii) the participant was determined to be cognitively normal based on thorough review of clinical and neuropsychological data at consensus diagnostic conference. MCI and dementia diagnoses were determined according to Petersen [19] and Diagnostic and Statistical Manual of Mental Disorders III-R criteria [20], respectively.

### 2.3 PET image acquisition and processing

Dynamic amyloid PET scans were acquired using ^11^C-PiB over 70 min on either a General Electric Advance scanner or a Siemens High Resolution Research Tomograph immediately following an intravenous bolus injection of approximately 555 MBq of radiotracer. Distribution volume ratio (DVR) was calculated using a spatially constrained simplified reference tissue model with a cerebellar gray matter reference region [21]. Mean cortical amyloid burden was calculated as the average DVR in the cingulate, frontal, parietal (including precuneus), lateral temporal, and lateral occipital regions, excluding the pre- and post-central gyri. Mean cortical DVR (cDVR) values were harmonized between the two scanners by leveraging longitudinal data available on both scanners for 79 participants. PET acquisition and processing are described in [22,23]. The number of longitudinal PiB PET measurements included was 589.

#### 2.3.1 PiB group determination

PiB PET scans were categorized as −/+ based on a cDVR threshold of 1.06 derived from a Gaussian mixture model fitted to harmonized cDVR values at first PET. We imputed PiB group for visits without a PiB PET scan (Supplementary Material).

### 2.4 Plasma biomarkers

Aβ_40_, Aβ_42_, GFAP, and NfL were measured at Johns Hopkins University (Baltimore, Maryland, USA) on a Quanterix (Billerica, Massachusetts, USA) HD-X instrument using the Quanterix Simoa Neurology 4-plex-E assay in duplicate and averaged (intra-assay coefficient of variation was 2.8, 1.9, 5.0, and 5.1, respectively [24]). Three outlying NfL measurements >125 pg/mL were excluded based on examination of within-individual longitudinal data. p-tau181 and p-tau231 were measured at the Clinical Neurochemistry Laboratory, University of Gothenburg (Mölndal, Sweden) on a Quanterix HD-X instrument using Simoa assays developed in-house [25,26]. Repeatability coefficients were 5.1% and 5.5% for the p-tau181 assay at concentrations of 11.6 and 15.5 pg/mL, respectively. Repeatability coefficients were 3.4% and 7.4% for the p-tau231 assay at concentrations of 31.6 and 42.7 pg/mL, respectively. For p-tau, concentrations below limit of detection were imputed at 0 and values below lower limit of quantitation were retained as is. In the main paper, we focus on the ratios Aβ_42_/Aβ_40_, p-tau181/Aβ_42_, and p-tau231/Aβ_42_ in addition to the concentrations of GFAP and NfL, and report results for the individual proteins Aβ_40_, Aβ_42_, p-tau181, and p-tau231 in the Supplementary Material. We divided p-tau concentrations by Aβ_42_ based on the performance of CSF p-tau181/Aβ_42_ in discriminating between PiB+ and PiB– [27,28] as well as other amyloid PET tracer-based positivity definitions [29] and in predicting conversion from a CDR of 0 to >0 [30]. Since reduction in CSF or plasma Aβ_42_ rather than Aβ_40_ is a better indicator of AD [31], dividing by Aβ_42_ yields a ratio more specific to AD. Plasma p-tau/Aβ_42_ is also associated with amyloid and tau PET [32,33]. The number of longitudinal measurements included across 199 participants was 685 for Aβ_40_, Aβ_42_ and GFAP, 682 for NfL, 671 for p-tau181, 676 for p-tau231, 597 for p-tau181/Aβ_42_, and 602 for p-tau231/Aβ_42_.

We estimated glomerular filtration rate (eGFR) at each plasma visit from serum creatinine levels using the Chronic Kidney Disease-Epidemiology collaboration formula. For visits without serum creatinine measurements, we imputed eGFR by carrying it forward or backward in time within person.

### 2.5 Statistical analysis

#### 2.5.1 Classification of brain amyloid status using plasma biomarkers

We assessed the performance of each plasma measure in classifying individuals into PiB groups at the index visit. We examined the receiver operating characteristic (ROC) curve and the area under the curve (AUC) separately for each measure. We also assessed the performance of plasma measures and demographics (age, sex, race, and *APOE ε*4 genotype) in multivariable analyses for classifying PiB group. As multivariable analyses involved estimating model parameters, we used 10-fold stratified (i.e., the proportion of PiB+ individuals in each fold was approximately the same) cross-validation to obtain ROC curves by estimating model parameters in the training set and obtaining predictions in the testing set. The models investigated included elastic net logistic regression models (with varying levels of ℓ_1_ and ℓ_2_ penalties to span the spectrum from Lasso to ridge regression), distributed random forests, gradient boosting machines, and extreme gradient boosting (XGBoost). Multivariable classifiers were fitted using automl in the H2O package (version 3.36.0.3) [34,35] in R version 4.0.3 [36].

#### 2.5.2 Longitudinal intraclass correlation coefficients

To assess the longitudinal reliability of biomarkers after accounting for expected population-level changes, we computed longitudinal intraclass correlation coefficients (ICC) using a linear mixed effects model (LMEM) for each biomarker that included an intercept and time from index visit term as fixed effects and a random intercept per participant. ICC was calculated as the ratio of the variance of the random intercept to the sum of the variances of the random intercept and noise. We calculated longitudinal ICC using data for (i) all, (ii) only PiB−, and (iii) only PiB+ individuals.

#### 2.5.3 Longitudinal plasma biomarker trajectories by brain amyloid status

We examined longitudinal plasma biomarker trajectories by brain amyloid status using a separate LMEM per biomarker. Unadjusted models included PiB group at index visit, time from index visit, and their interaction. Adjusted models additionally included age at index visit, sex, race, *APOE ε*4 status, and age × time interaction. We also included eGFR and body mass index (BMI) concurrent with plasma measurement as covariates given their associations with plasma biomarkers [37]. The main goals of this analysis were to examine PiB group differences in (i) plasma concentrations at index visit and (ii) longitudinal rates of change in plasma concentrations for each of the five measures: Aβ_42_/Aβ_40_, p-tau181/ Aβ_42_, p-tau231/Aβ_42_, GFAP, and NfL. Statistical significance was defined as two-tailed *p* < 0.01. This threshold is based on Bonferroni correction to achieve a 5% family-wise error rate based on five hypothesis tests in each family of hypotheses. In addition to examining PiB group differences, we conducted post-hoc analyses to examine slope within each PiB group, but we do not report these in the main text unless the PiB group × time interaction was statistically significant.

#### 2.5.4 Associations among longitudinal rates of change in plasma biomarkers and brain amyloid

We used bivariate LMEMs to examine the association between rates of change in pairs of biomarkers. We considered longitudinal data for two biomarkers simultaneously as dependent variables. Independent variables were age at index visit, time from index visit, age × time interaction, sex, race, and *APOE ε*4 status. For plasma biomarkers, we additionally adjusted for eGFR and BMI concurrent with plasma measurement. We estimated a separate noise variance per outcome. We included a random intercept and slope over time per participant for each outcome. The covariance of these four random effects was modeled using an unstructured covariance matrix, from which we extracted the correlation between random slopes to assess the association between rates of biomarker change. Bivariate LMEMs were fitted using the lme function and correlation parameter confidence intervals were computed using the intervals function in the nlme package [38]. Statistical significance was defined as two-tailed *p* < 0.0033. This threshold is based on Bonferroni correction to achieve a 5% family-wise error rate based on 15 hypothesis tests (one for each pair among six biomarkers, including five plasma biomarkers in the main analysis and one PiB PET measure).

#### 2.5.5 Temporal order of changes in plasma biomarkers and brain amyloid

We assessed the temporal order of changes in plasma biomarkers and cDVR using a Bayesian implementation of the progression score (PS) model (modified from [39]). The PS model accounts for individual differences in the onset of biomarker changes by estimating a time-shift per individual to better align longitudinal measurements. We modeled biomarker trajectories using sigmoid functions. This analysis was limited to 577 longitudinal visits across 199 participants where the full set of plasma biomarkers and cDVR were available.

To confirm that PS reflects disease progression, we assessed whether PS at last visit and the time-shift variable *τ* were higher among individuals with MCI or dementia compared to cognitively normal individuals. Since cognitive diagnosis is not used in the fitting of the PS model, this variable provides an independent way of validating the PS.

## 3 Results

### 3.1 Descriptives

Participant demographics are presented in Table 1. Compared to PiB−, PiB+ individuals were more likely to be *APOE ε*4 +, had lower plasma Aβ_42_/Aβ_40_, higher Aβ_40_, p-tau181, p-tau231, p-tau181/Aβ_42_, p-tau231/Aβ_42_, GFAP, and NfL at index visit, and were less likely to remain cognitively normal. At index visit, eGFR was positively correlated with Aβ_42_/Aβ_40_ (*r* = 0.18, 95% CI = [0.039, 0.31], *p* = 0.013) and negatively correlated with the remaining plasma measures (*τ* ranging from -0.45 to -0.17, all *p* < 0.018). BMI was negatively correlated with p-tau181/Aβ_42_ (*r* = -0.14, 95% CI = [-0.28, -0.0044], *p* = 0.043), GFAP (*r* = -0.28, 95% CI = [-0.41, -0.15], *p* = 5.17 × 10^-5^), and NfL (*r* = -0.27, 95% CI = [-0.4, -0.14], *p* = 8.60 × 10^-5^). Men had lower Aβ_42_/Aβ_40_ and higher Aβ_40_, p-tau181, p-tau231, p-tau181/Aβ_42_, p-tau231/Aβ_42_, GFAP, and NfL compared to women. White participants had higher p-tau181, p-tau181/Aβ_42_, GFAP, and NfL compared to Non-White participants. Relationships of plasma and PiB PET measures with eGFR, BMI, sex, and race are shown in Supplementary Figures 1–4. We did not observe associations of eGFR, BMI, sex, or race with PiB cDVR. Correlations among plasma and PiB PET measures at index visit are presented in Supplementary Figure 5 and longitudinal measures versus age in Supplementary Figure 6.

**Table 1:** Participant characteristics. Time-varying variables are summarized at the index visit. For continuous and categorial variables, we report the median and interquartile range and the N and percentage, respectively. PiB group comparisons are based on Wilcoxon rank-sum test for continuous variables and Pearson’s Chi-squared test or Fisher’s exact test for categorical variables.

| Characteristic | Overall, N = 199 | PiB-, N = 141 | PiB+, N = 58 | p-value |
| --- | --- | --- | --- | --- |
| Age (yr) | 76 (69, 82) | 74 (68, 81) | 79 (73, 84) | 0.005 |
| Male | 97 (49%) | 66 (47%) | 31 (53%) | 0.4 |
| Race |  |  |  | >0.9 |
| API and Other | 9 (4.5%) | 7 (5.0%) | 2 (3.4%) |  |
| Black | 34 (17%) | 25 (18%) | 9 (16%) |  |
| White | 156 (78%) | 109 (77%) | 47 (81%) |  |
| APOE ε4+ | 59 (30%) | 36 (26%) | 23 (40%) | 0.047 |
| eGFR (mL/min/1.73 m<sup>2</sup>) | 73 (63, 85) | 74 (64, 86) | 72 (59, 79) | 0.079 |
| BMI (kg/m<sup>2</sup>) | 26.7 (24.2, 30.1) | 27.4 (24.2, 31.1) | 26.3 (24.2, 29.0) | 0.2 |
| Aβ<sub>40</sub> (pg/mL) | 139 (114, 169) | 133 (111, 163) | 150 (123, 184) | 0.011 |
| Aβ<sub>42</sub> (pg/mL) | 6.95 (5.62, 8.16) | 7.24 (5.67, 8.17) | 6.48 (5.51, 7.84) | 0.2 |
| p-tau181 (pg/mL) | 7 (5, 11) | 7 (5, 9) | 11 (8, 18) | <0.001 |
| p-tau231 (pg/mL) | 18 (13, 24) | 16 (13, 20) | 27 (16, 36) | <0.001 |
| Aβ<sub>42</sub>/Aβ<sub>40</sub> | 0.050 (0.044, 0.056) | 0.052 (0.047, 0.060) | 0.046 (0.040, 0.051) | <0.001 |
| p-tau181/Aβ<sub>42</sub> | 1.11 (0.77, 1.72) | 0.93 (0.70, 1.43) | 1.71 (1.28, 2.89) | <0.001 |
| p-tau231/Aβ<sub>42</sub> | 2.51 (1.87, 3.56) | 2.22 (1.81, 2.99) | 4.05 (2.62, 5.94) | <0.001 |
| GFAP (pg/mL) | 185 (131, 251) | 173 (122, 217) | 229 (185, 301) | <0.001 |
| NfL (pg/mL) | 23 (17, 31) | 22 (16, 28) | 27 (20, 36) | 0.002 |
| Hypertension | 107 (54%) | 83 (59%) | 24 (41%) | 0.025 |
| Diabetes | 38 (19%) | 26 (18%) | 12 (21%) | 0.7 |
| High cholesterol | 114 (57%) | 84 (60%) | 30 (52%) | 0.3 |
| Obesity | 53 (27%) | 44 (31%) | 9 (16%) | 0.023 |
| Smoking |  |  |  | 0.5 |
| Never | 101 (52%) | 73 (53%) | 28 (50%) |  |
| Former | 90 (46%) | 62 (45%) | 28 (50%) |  |
| Current | 4 (2.1%) | 4 (2.9%) | 0 (0%) |  |
| Unknown | 4 | 2 | 2 |  |
| Number of visits | 3 (2, 5) | 3 (2, 5) | 4 (2, 5) | 0.8 |
| Follow-up duration (yr) | 6.1 (4.0, 8.6) | 6.1 (4.0, 8.6) | 6.1 (3.8, 8.7) | 0.6 |
| Final diagnosis |  |  |  | 0.001 |
| Cognitively normal | 157 (79%) | 121 (86%) | 36 (62%) |  |
| MCI | 20 (10%) | 9 (6.4%) | 11 (19%) |  |
| Other impairment | 1 (0.5%) | 0 (0%) | 1 (1.7%) |  |
| Dementia | 21 (11%) | 11 (7.8%) | 10 (17%) |  |

Abbreviations: Aβ , amyloid-beta; API, Asian/Pacific Islander; APOE, apolipoprotein E; BMI, body mass index; eGFR, estimated glomerular filtration rate; GFAP, glial fibrillary acidic protein; MCI, mild cognitive impairment; NfL, neurofilament light chain; PiB, Pittsburgh compound B; p-tau, phosphorylated tau.

### 3.2 Classification of brain amyloid status using plasma biomarkers

#### 3.2.1 Univariate models based on a single plasma biomarker or biomarker ratio

ROC curves for univariate models are presented in Figure 1 and Supplementary Figure 7. The best univariate classifiers were p-tau231/Aβ_42_, p-tau181/Aβ_42_, and p-tau231, with AUCs in the range 0.76–0.78 (Supplementary Table 1). The performance of the NfL-only classifier (AUC = 0.64, 95% CI = [0.55–0.72]) was similar to that of the age-only classifier (AUC = 0.63, 95% CI = [0.54–0.71]), whereas Aβ_42_/Aβ_40_ (AUC = 0.72, 95% CI = [0.65–0.79]), p-tau181 (AUC = 0.72, 95% CI = [0.63–0.8]), p-tau231 (AUC = 0.76, 95% CI = [0.67–0.85]), p-tau181/Aβ_42_ (AUC = 0.77, 95% CI = [0.7–0.84]), p-tau231/Aβ_42_ (AUC = 0.78, 95% CI = [0.71–0.86]), and GFAP (AUC = 0.71, 95% CI = [0.63–0.79]) outperformed age.

**Figure 1:**
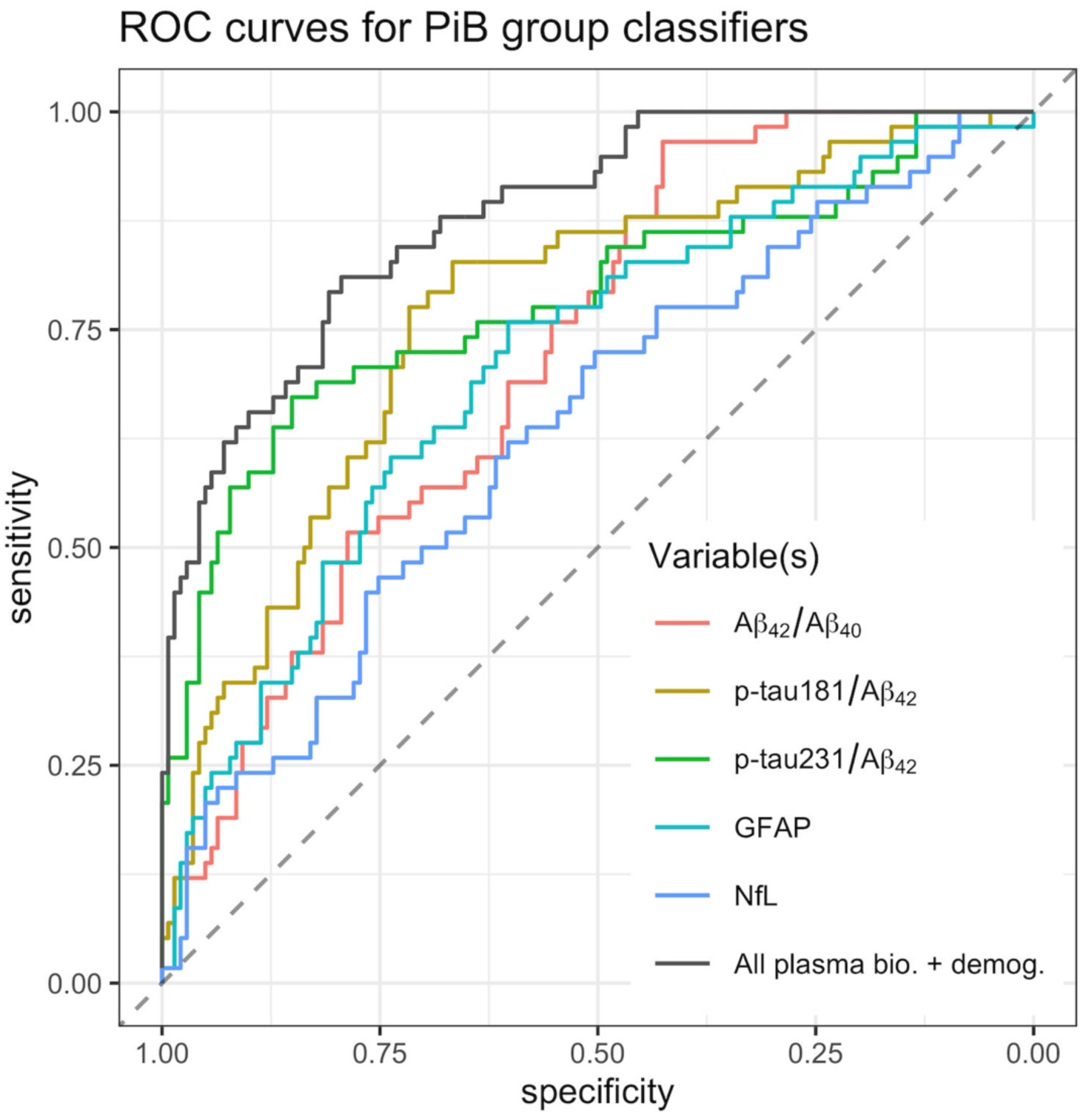
Receiver operating characteristic curves for univariate classifiers and the best multivariable classifier (including all plasma measures, age, sex, race, and APOE ε4 status as features) for predicting PiB group. A*β*, amyloid-*β*; APOE, apolipoprotein E; bio., biomarkers; demog., demographics; GFAP, glial fibrillary acidic protein; NfL, neurofilament light chain; PiB, Pittsburgh compound B; p-tau, phosphorylated tau; ROC, receiver operating characteristic.

#### 3.2.2 Multivariable models

Classifiers based on multiple predictors had slightly better performance than classifiers based on single predictors. The highest AUC classifier was a gradient boosting machine, yielding an AUC = 0.88 (95% CI = [0.73, 0.89]). At the operating point with the highest balanced accuracy, this classifier achieved 79% specificity and 81% sensitivity (Figure 1). This classifier outperformed the best demographics-only multivariate classifier (stacked ensemble with AUC = 0.70).

To identify the most parsimonious model, we first calculated feature importance from the best gradient boosting machine classifier. Variables with the highest importance were p-tau231 and Aβ_42_/Aβ_40_. A gradient boosting machine classifier with these two variables yielded an AUC = 0.89, suggesting that this model with only two plasma measures achieves a PiB group classification performance comparable to that of the model with all demographics and plasma measures.

### 3.3 Longitudinal intraclass correlation coefficients

Longitudinal ICCs over a median follow-up of 6.1 years (IQR: 4, 8.6) are presented in Table 2 and Supplementary Table 2. Plasma measures had lower longitudinal ICC than that of cDVR in the whole sample and among PiB+ individuals, suggesting that their longitudinal rates of change are not as reliable as that of cDVR.

**Table 2:** Longitudinal intraclass correlation coefficients (ICCs).

| Biomarker | Overall |  | PiB– |  | PiB+ |  |
| --- | --- | --- | --- | --- | --- | --- |
|  | ICC | 95% CI | ICC | 95% CI | ICC | 95% CI |
| Aβ<sub>42</sub>/Aβ<sub>40</sub> | 0.66 | (0.6–0.72) | 0.67 | (0.57–0.75) | 0.68 | (0.56–0.78) |
| p-tau181/Aβ<sub>42</sub> | 0.67 | (0.59–0.73) | 0.61 | (0.5–0.7) | 0.62 | (0.47–0.73) |
| p-tau231/Aβ<sub>42</sub> | 0.75 | (0.69–0.8) | 0.57 | (0.46–0.66) | 0.75 | (0.63–0.83) |
| GFAP | 0.78 | (0.72–0.82) | 0.79 | (0.73–0.84) | 0.77 | (0.67–0.85) |
| NfL | 0.67 | (0.6–0.72) | 0.72 | (0.64–0.79) | 0.63 | (0.48–0.74) |
| PiB cDVR | 0.96 | (0.94–0.97) | 0.70 | (0.62–0.77) | 0.96 | (0.94–0.98) |

Abbreviations: Aβ , amyloid-beta; cDVR, cortical distribution volume ratio; CI, confidence interval; GFAP, glial fibrillary acidic protein; ICC, intraclass correlation coefficient; NfL, neurofilament light chain; PiB, Pittsburgh compound B; p-tau, phosphorylated tau.

### 3.4 Longitudinal plasma biomarker trajectories by brain amyloid status

At the index visit, PiB+ individuals had lower Aβ_42_/Aβ_40_ (*β* = −7.58 × 10^-3^, SE = 1.41 × 10^-3^, *p* = 2.36 × 10^-7^) and higher p-tau181/Aβ_42_ (*β* = 0.599, SE = 0.129, *p* = 6.16 × 10^-6^), p-tau231/Aβ_42_ (*β* = 1.86, SE = 0.243, *p* = 1.28 × 10^-12^), and GFAP (*β* = 44.1, SE = 11.6, *p* = 1.81 × 10^-4^) in adjusted models (Figure 2 and Supplementary Table 3). PiB groups exhibited different rates of longitudinal change in Aβ_42_/Aβ_40_ (PiB group × time interaction *β* = 5.41 × 10^-4^, SE =1.95 × 10^-4^, *p* = 0.0073); post-hoc analyses showed that rate of change was not statistically significant among PiB+ individuals while PiB− individuals exhibited decreases (*β* = −3.85 × 10^-4^, SE = 9.77 × 10^-5^, *p* = 1.96 × 10^-4^) (Supplementary Table 4). We did not find statistically significant PiB group differences in rates of change for the remaining plasma measures in adjusted models.

**Figure 2:**
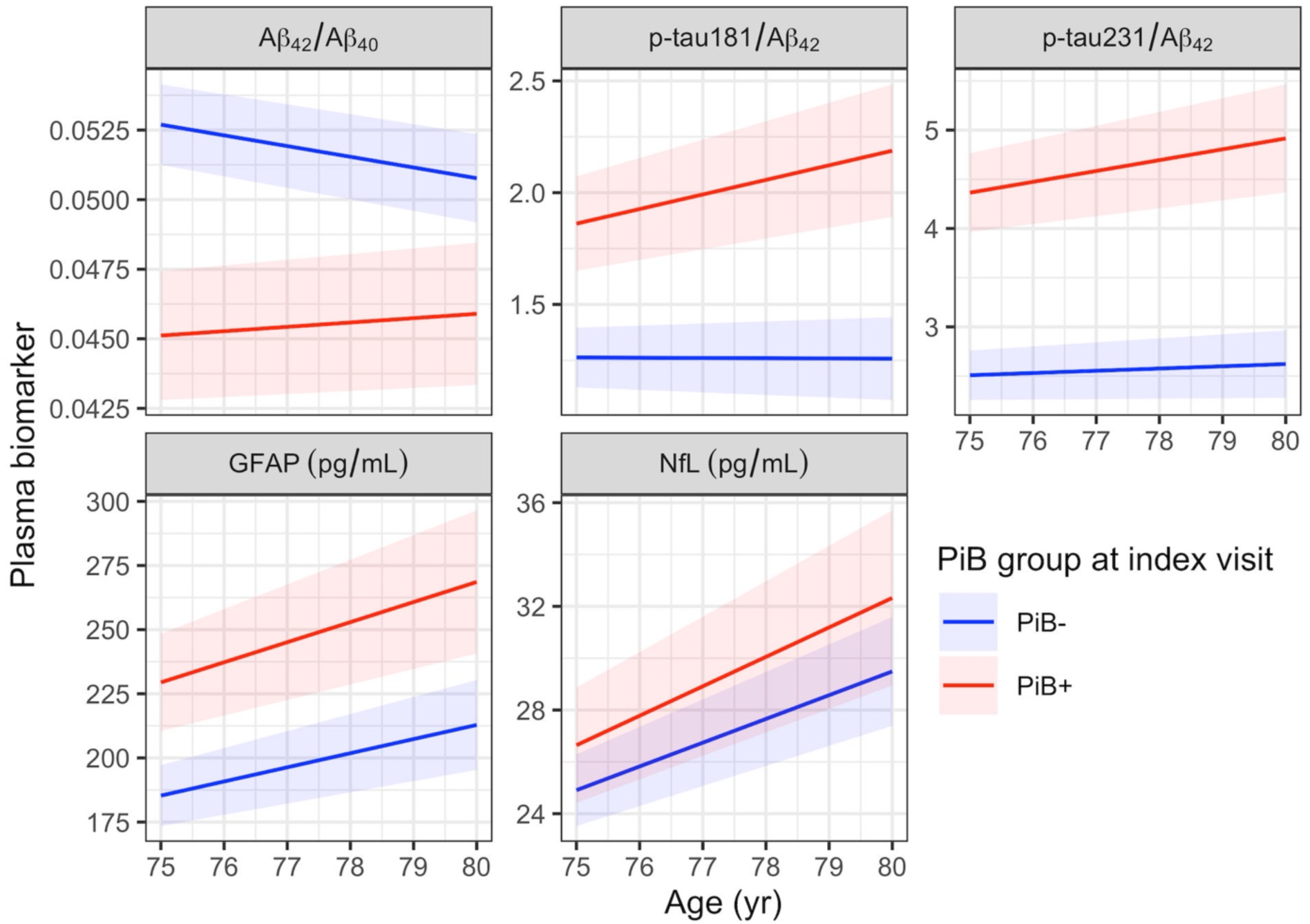
Plasma biomarker trajectories estimated using linear mixed effects models. A linear mixed effects model was fitted per biomarker. Models included PiB group at index visit, time from index visit, and their interaction, allowing for the calculation of an average biomarker trajectory per PiB group. Models additionally adjusted for age at index visit, sex, race, APOE ε4 status, and age × time interaction. Bands indicate 95% confidence intervals. A*β*, amyloid-*β*; GFAP, glial fibrillary acidic protein; NfL, neurofilament light chain; PiB, Pittsburgh compound B; p-tau, phosphorylated tau.

### 3.5 Associations among longitudinal rates of change in plasma biomarkers and brain amyloid

The correlation between longitudinal rates of change in p-tau181/Aβ_42_ and p-tau231/Aβ_42_ was high and statistically significant (*r* = 0.87, 95% CI = [0.62, 0.96], *p* < 0.001) (Supplementary Figure 9). We additionally found statistically significant correlations between the rates of change in GFAP and NfL (*r* = 0.88 [0.63, 0.97], *p* < 0.001) and GFAP and cDVR (*r* = 0.5 [0.26, 0.68], *p* < 0.001). The correlation between rates of change in NfL and cDVR (*r* = 0.4 [0.13, 0.62], *p* = 0.0043) did not survive multiple comparison correction.

### 3.6 Temporal order of changes in plasma biomarkers and brain amyloid

Estimated PS and biomarker trajectories, along with observed biomarker data, are shown in Figure 3. Consistent with expectation, both PS at last visit and the subject-specific time-shift parameter were higher among individuals with MCI or dementia compared to cognitively normal individuals (Wilcoxon rank-sum test *p* = 4.51 × 10^-6^ for PS, *p* = 0.0032 for time-shift variable *τ*).

**Figure 3:**
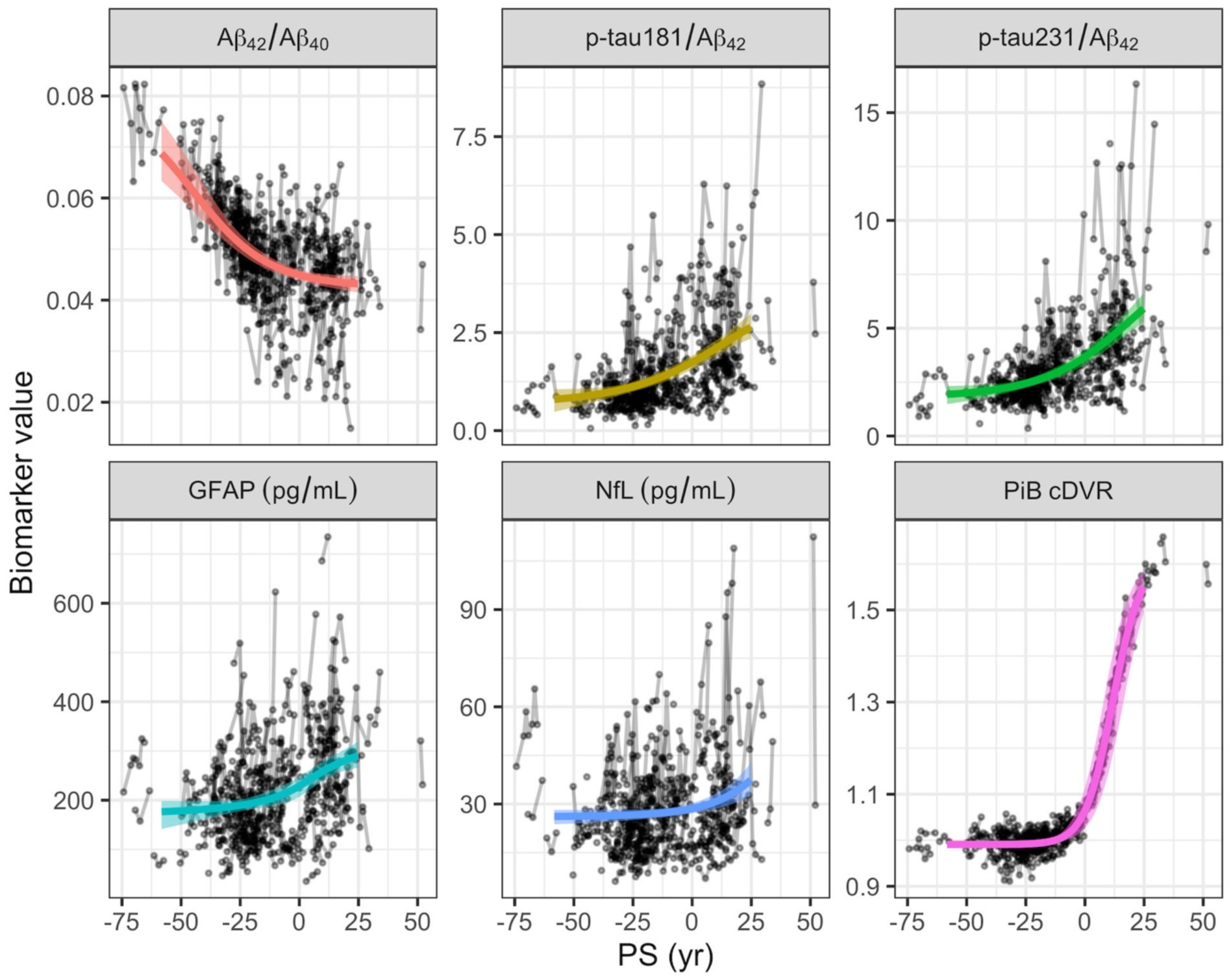
Biomarker trajectories estimated after alignment of individual-level longitudinal data using the progression score (PS) model. Bands indicate the 95% confidence intervals for the trajectory estimates. PS scale was calibrated after model fitting such that at PS = 0, the estimated trajectory for PiB cDVR attains the value 1.06, which is the PiB positivity threshold. Since PS is time-shifted age, it is in the units of years. A*β*, amyloid-*β*; cDVR, cortical distribution volume ratio; GFAP, glial fibrillary acidic protein; NfL, neurofilament light chain; PiB, Pittsburgh compound B; PS, progression score; p-tau, phosphorylated tau.

To understand the relative order of biomarker changes, we computed percent relative change by dividing the derivative in PS of the estimated trajectory by the trajectory itself for each biomarker (Figure 4 and Supplementary Figure 10) and examined where the peak percent relative change occurs relative to the PS value corresponding to the PiB+ threshold. This analysis suggested that the earliest change occurs in Aβ_42_/Aβ_40_. Peak relative decline in Aβ_42_/Aβ_40_ (-1% per year) preceded brain amyloid positivity onset by 41 years (95% CI = [32, 53]) (Supplementary Table 5). Time intervals between brain amyloid positivity onset and peak relative change in the remaining plasma biomarkers were not statistically significant.

**Figure 4:**
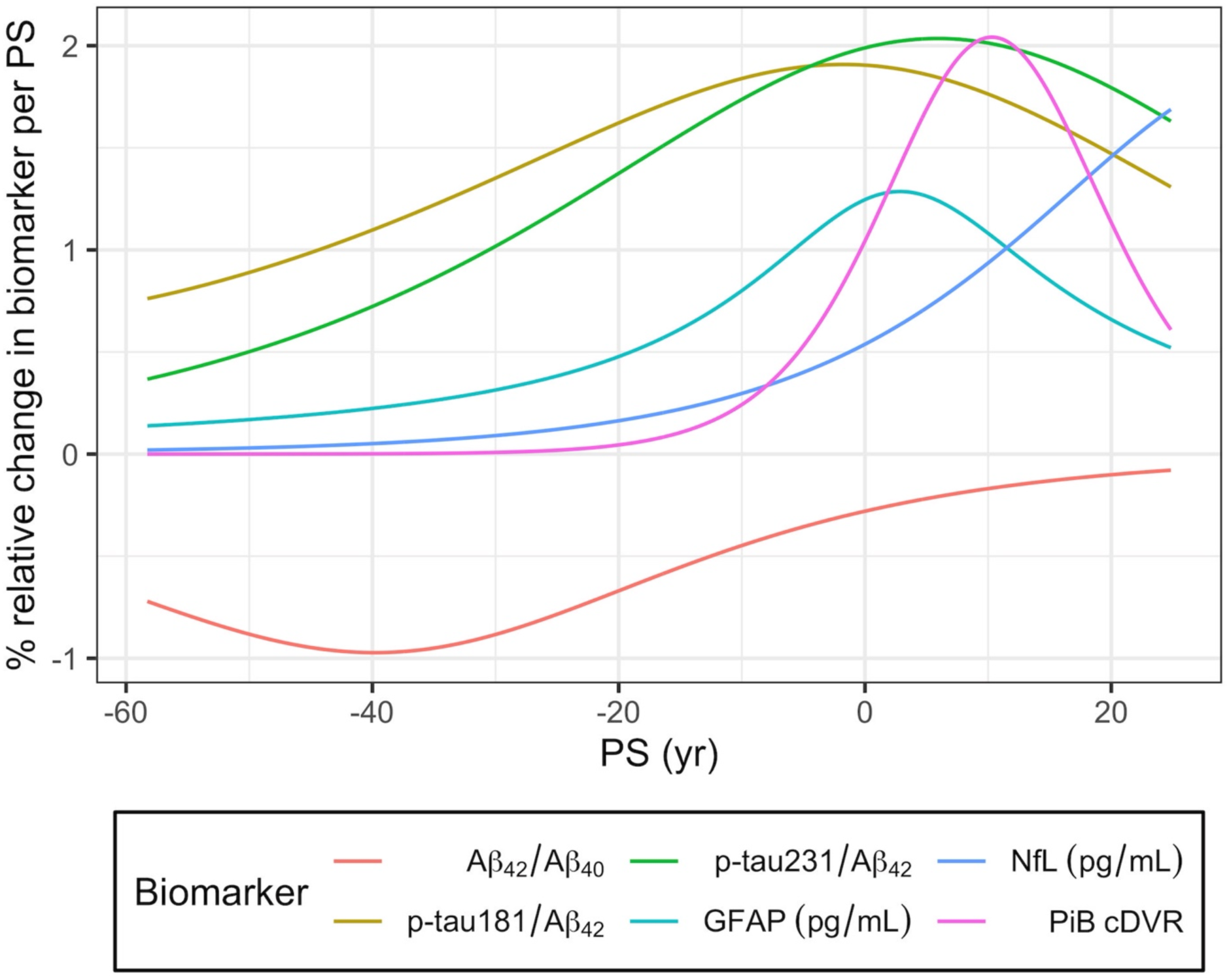
Percent relative change in biomarkers per progression score (PS) as a function of PS. A*β*, amyloid-*β*; cDVR, cortical distribution volume ratio; GFAP, glial fibrillary acidic protein; NfL, neurofilament light chain; PiB, Pittsburgh compound B; PS, progression score; p-tau, phosphorylated tau.

## 4 Discussion

This study focused on longitudinal changes in plasma biomarkers of AD neuropathology and neurodegeneration relative to amyloid plaques, the emergence of which marks the beginning of preclinical AD. We first replicated prior findings of the extent to which plasma biomarkers predict PET brain amyloid status. In our sample of cognitively normal individuals, the plasma measures with the best amyloid PET status classification performance were the p-tau to Aβ_42_ ratios. Our AUCs based on single plasma biomarkers are consistent with AUCs reported in other studies of cognitively normal individuals based on Simoa immunoassays [15,40–42]. Our findings also corroborate previous studies indicating that plasma p-tau measures more closely reflect brain amyloid levels compared to plasma measures of amyloid [16] and that p-tau231 has the highest AUC at the preclinical stage [14–16]. As expected based on our univariate results, plasma p-tau, specifically p-tau231, and Aβ measures were the most important variables in the best multivariable classifier, which outperformed univariate classifiers and had a sensitivity and specificity of about 80% at its optimal operating point.

The main contribution of our paper is the longitudinal examination of changes in plasma biomarkers. Longitudinal reliability, as measured by ICC, of plasma measures was lower than that of the brain amyloid PET measure in the whole sample and among PiB+, but comparable among PiB−. Longitudinal decrease in plasma Aβ_42_/Aβ_40_ was statistically significant among PiB− individuals, but not among PiB+. This, along with the finding that PiB+ individuals have lower Aβ_42_/Aβ_40_ at index visit compared to PiB−, suggests that plasma Aβ_42_/Aβ_40_ declines prior to the emergence of elevated levels of brain amyloid and then may reach a plateau. Other studies have also found that amyloid PET is elevated or increases only when plasma Aβ_42_/Aβ_40_ is low [43,44].

The plasma measure that most closely changed in conjunction with brain amyloid levels was GFAP. Rates of change in NfL also aligned with rate of change in brain amyloid level. Plasma Aβ_42_/Aβ_40_ did not correlate longitudinally with brain amyloid or any other plasma biomarker. This difference in the longitudinal correlations for brain and plasma amyloid is likely due to the different time windows in which these two measures are dynamic, with plasma amyloid exhibiting changes decades prior to brain amyloid. Our findings agree with the plasma biomarker findings from the TRAILBLAZER-ALZ clinical trial, where longitudinal change in brain amyloid correlated with change in plasma GFAP but not Aβ_42_/Aβ_40_ [45].

Our progression score model suggests that Aβ_42_/Aβ_40_ may decline over several decades leading up to the onset of brain amyloid accumulation. However, these changes in plasma Aβ_42_/Aβ_40_ are subtle, with relative change peaking at -1% per year. Brain PET measures fibrillar amyloid, an advanced stage in the amyloid aggregation process, whereas plasma biomarkers reflect earlier soluble forms [46]. This difference is one possible explanation of the timing difference between plasma and brain amyloid measures. These results suggest that if it can be measured with high accuracy and longitudinal reliability, plasma Aβ_42_/Aβ_40_ may allow for detecting early changes prior to the emergence of brain amyloid plaques.

Given that plasma Aβ_42_/Aβ_40_may plateau by the time one has high levels of brain amyloid, its utility in a longitudinal context among amyloid PET positive individuals is likely limited. Other plasma biomarkers we investigated exhibited more pronounced changes over time, with p-tau ratios exhibiting relative changes around 2% per year, and these changes occurred closer in time to brain amyloid accumulation. This finding is consistent with literature demonstrating that plasma p-tau measurements better align with brain amyloid rather than tau levels as measured with PET [47,48]. Our results regarding longitudinal changes and temporal order are consistent with other studies that investigated longitudinal plasma measurements [10–12]. More extensive longitudinal data will allow examination of temporal order variation at the individual level.

Our study has several limitations. More recent measures of plasma Aβ exhibit stronger associations with brain amyloid compared to the Quanterix Simoa measure that we used [49]. It is possible that we were unable to detect a statistically significant PiB group difference in the longitudinal rates of change in p-tau, GFAP, and NfL due to the limited number of participants included in our study and the lower longitudinal ICC of plasma measures, in particular, the p-tau ratios. The characterization of biomarker trajectories was informed mainly by data from cognitively normal individuals, and the lack of data from late dementia stages prevented us from describing the full extent of the natural history of these biomarkers. The longitudinal follow-up duration was much shorter than the estimated time intervals over which plasma biomarkers change, preventing us from verifying our estimates using individual-level data. It will be important to validate these findings using independent samples with more individuals and longer follow-up.

Our study also has important strengths. The median follow-up duration for our plasma measures, 6.1 years, is higher than the follow-up duration of 2–4 years in existing longitudinal plasma biomarker studies [8–13]. We used advanced multivariable classifiers and employed cross-validation to calculate ROCs and classification performance metrics to prevent overestimating classifier performance. When investigating associations among rates of longitudinal change, instead of calculating slopes and then correlating them, we employed bivariate LMEMs, which factor in the uncertainty in the slopes when estimating correlations.

In conclusion, our results corroborate p-tau231 as a superior biomarker of amyloid burden in preclinical disease but suggest that plasma Aβ_42_/Aβ_40_ is dynamic prior to amyloid PET positivity. Other plasma measures, GFAP in particular, may more closely align with longitudinal change in brain amyloid accumulation. Plasma biomarkers are promising tools for detecting and monitoring longitudinal change along the disease spectrum and can help identify candidates for an amyloid PET scan. Given the emerging anti-amyloid therapies, assessing brain amyloid using easy and low cost measures such as plasma biomarkers will be particularly useful and important.

## Data Availability

BLSA data are available upon request from https://www.blsa.nih.gov. All requests are reviewed by the BLSA Data Sharing Proposal Review Committee.

https://gitlab.com/bilgelm/longitudinal-plasma-and-pib

