## Supplementary Material for "Longitudinal changes in Alzheimer’s-related plasma biomarkers and brain amyloid"

#### Sample inclusion and exclusion criteria

Of the 229 Baltimore Longitudinal Study of Aging (BLSA) participants with amyloid PET scans prior to March 13, 2020, one was excluded due to a previously unreported myocardial infarction prior to enrollment (an exclusion criterion), four were excluded due to insufficient or inadequate PET data that prevented image processing, and three were excluded due to missing apolipoprotein E (*APOE*) genotyping. Of the remaining 221 participants, 199 had at least one visit with measurements for the full set of plasma biomarkers. Data from these 199 participants comprised the current cross-sectional and longitudinal datasets.

The index visit, defined as the earliest cognitively normal visit with a full set of plasma biomarker measurements, overlapped with an amyloid PET visit for all except two participants. For these two participants, the index visit was prior to their PET scans. We imputed their PiB group at the index visit (see below). The resulting cross-sectional dataset was used to examine the performance of plasma biomarkers in classifying individuals as PiB− or PiB+.

In longitudinal analyses, we included all plasma measures following the index visit as well as all plasma measures preceding the index visit if the (imputed) PiB group at the preceding visit was the same as the (imputed) PiB group at the index visit.

#### PiB group imputation

In the longitudinal PiB PET dataset for the participants included in this analysis, no individuals reverted from PiB+ to PiB−. All visits prior to a PiB− scan were assumed to be PiB−, and all visits following a PiB+ scan were assumed to be PiB+. Visits within one year of a PiB PET scan were imputed using the binary PiB group assigned to the PiB PET scan. Otherwise, for visits preceding a PiB+ scan, we used the participant's estimated amyloid onset age [1] to impute PiB group. If the visit occurred prior to estimated amyloid onset, it was imputed as PiB−. If the visit occurred more than 3 years following the estimated amyloid onset, it was imputed as PiB+. Otherwise, we did not impute PiB group.

### Data and code availability

Code for performing statistical analyses and generating figures is provided in an open repository (https://gitlab.com/bilgelm/longitudinal-plasma-and-pib). BLSA data are available upon request from https://www.blsa.nih.gov. All requests are reviewed by the Data Sharing Proposal Review Committee.

#### Associations of plasma and PiB PET measures at index visit with eGFR, BMI, sex, and race

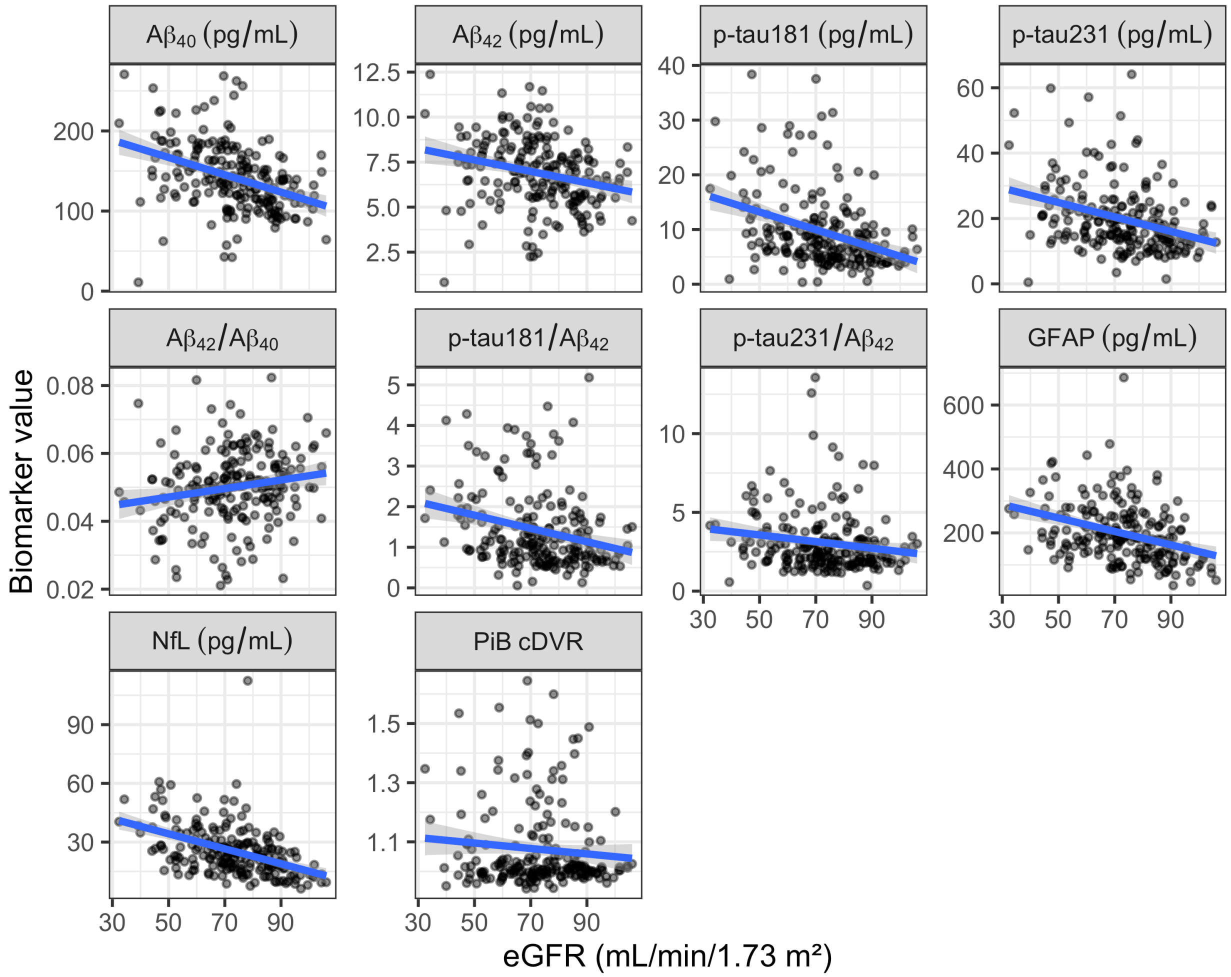


***Supplementary Figure 1: Plasma and PiB PET measures at the index visit as a function of estimated glomerular filtration rate (eGFR).*** *Aβ, amyloid-β; cDVR, cortical distribution volume ratio; GFAP, glial fibrillary acidic protein; NfL, neurofilament light chain; PiB, Pittsburgh compound B; p-tau, phosphorylated tau.*

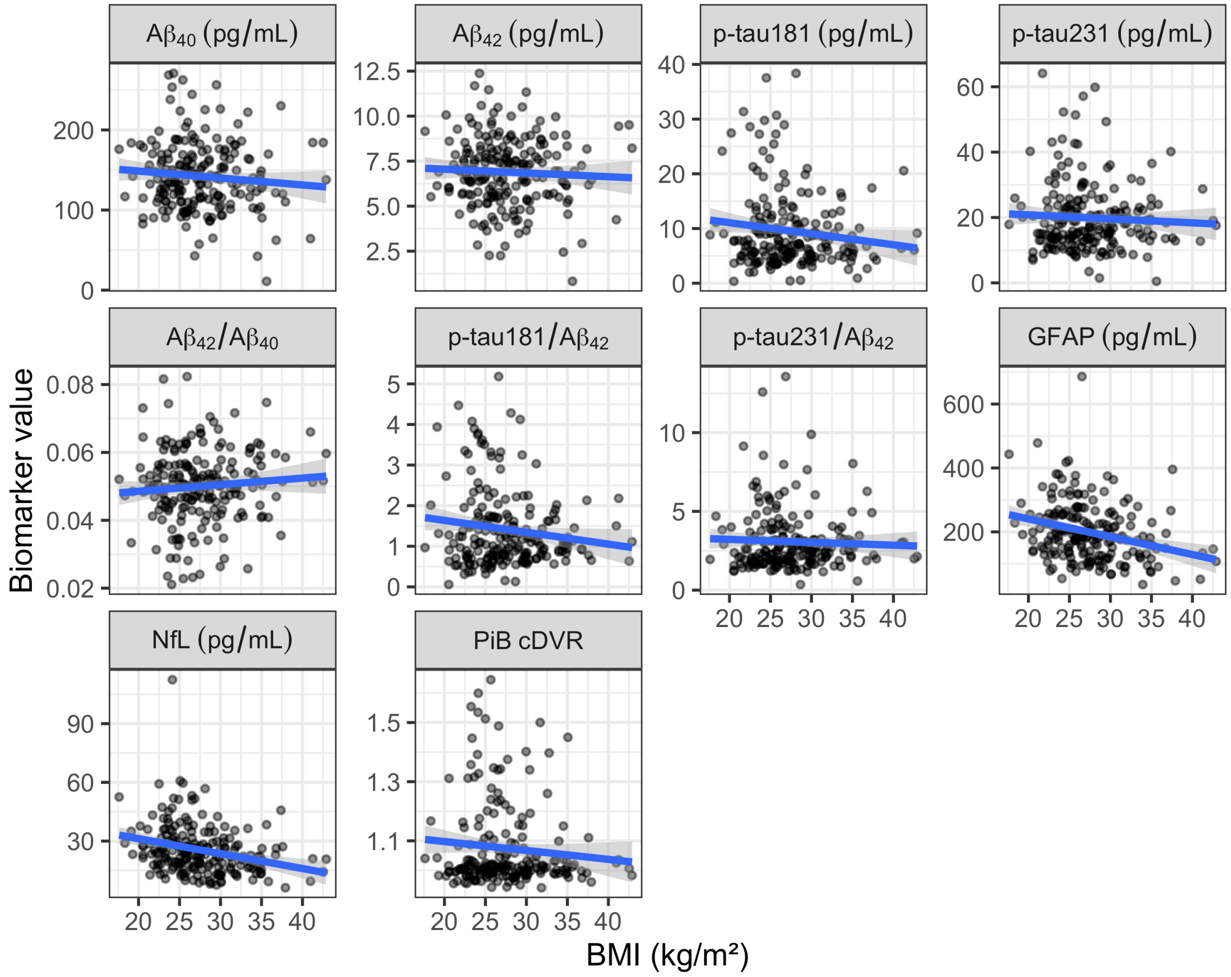


***Supplementary Figure 2: Plasma and PiB PET measures at the index visit as a function of body mass index (BMI).*** *Aβ, amyloid-β; cDVR, cortical distribution volume ratio; GFAP, glial fibrillary acidic protein; NfL, neurofilament light chain; PiB, Pittsburgh compound B; p-tau, phosphorylated tau.*

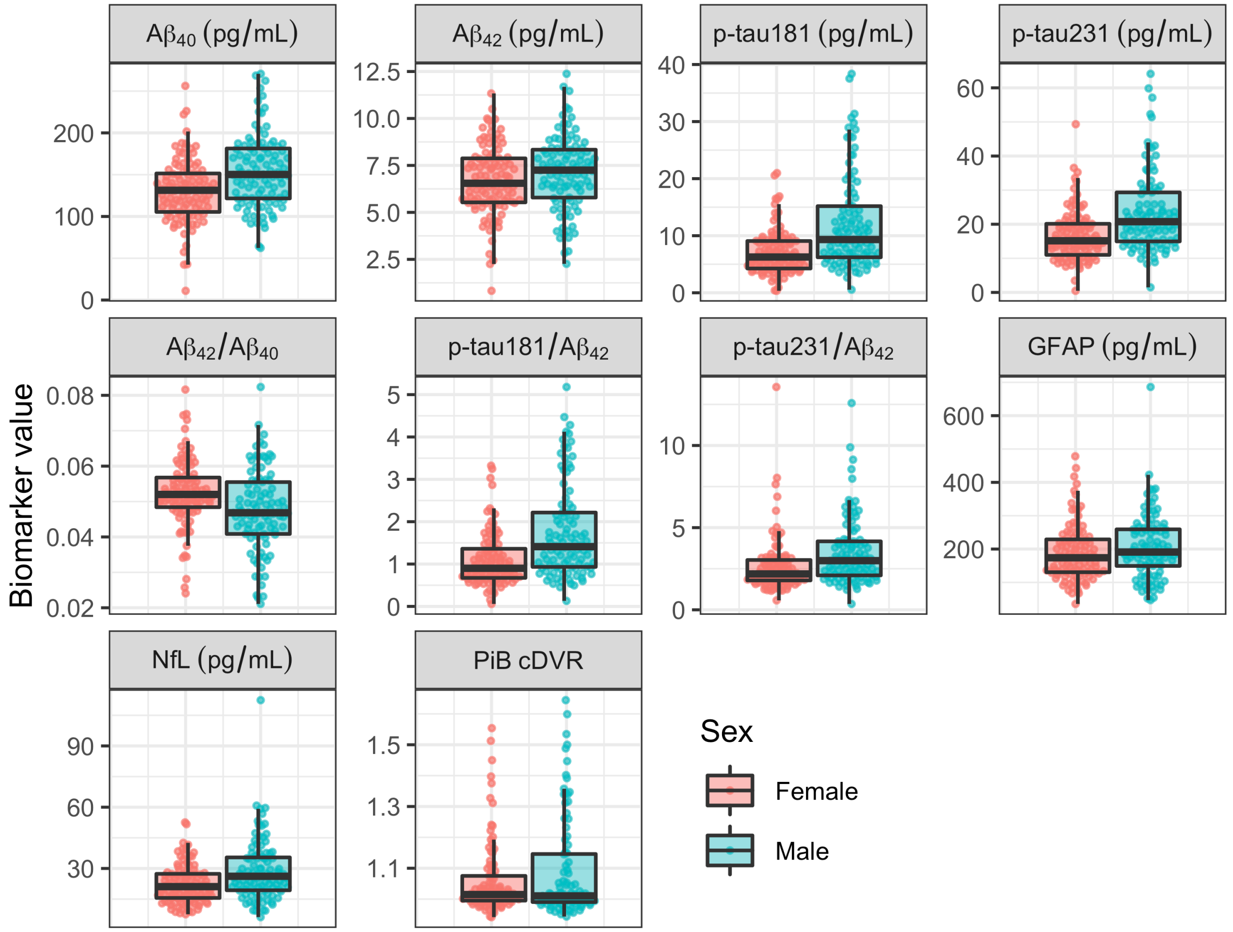


***Supplementary Figure 3: Plasma and PiB PET measures at the index visit by sex.*** *Aβ, amyloid-β; cDVR, cortical distribution volume ratio; GFAP, glial fibrillary acidic protein; NfL, neurofilament light chain; PiB, Pittsburgh compound B; p-tau, phosphorylated tau.*

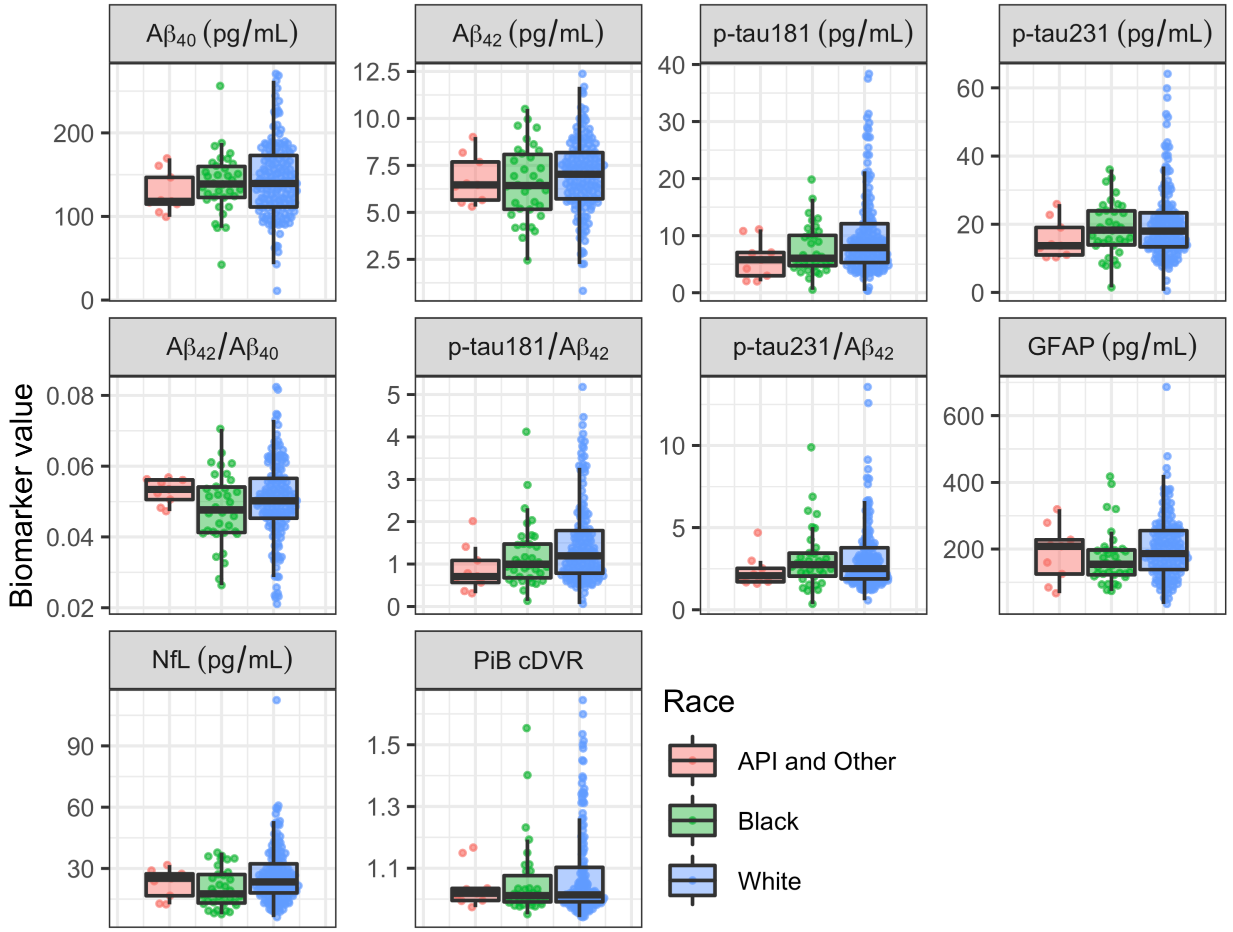


***Supplementary Figure 4: Plasma and PiB PET measures at the index visit by race.*** *Aβ, amyloid-β; API, Asian/Pacific Islander; cDVR, cortical distribution volume ratio; GFAP, glial fibrillary acidic protein; NfL, neurofilament light chain; PiB, Pittsburgh compound B; p-tau, phosphorylated tau.*

#### Correlations among plasma biomarkers and brain amyloid at index visit

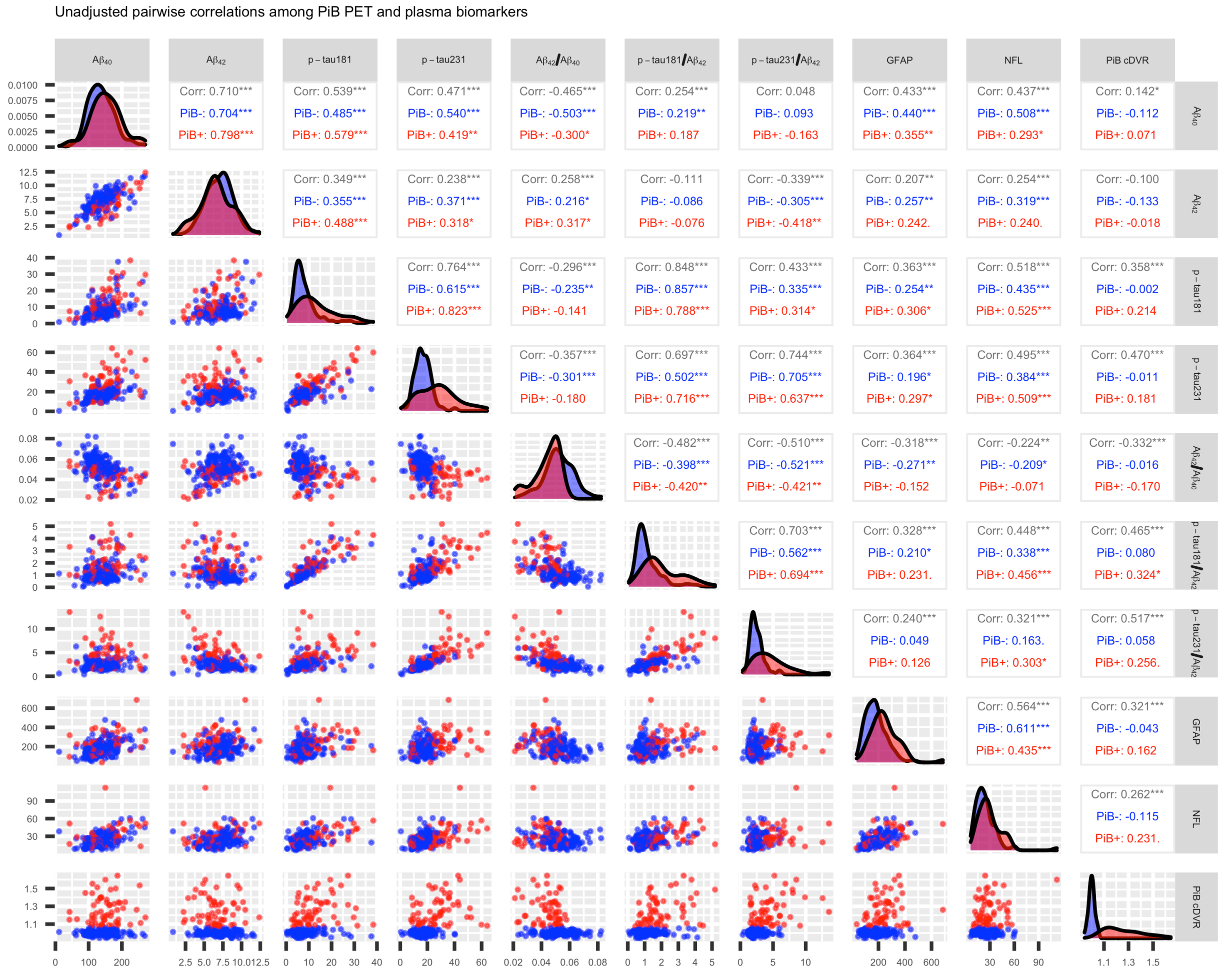


***Supplementary Figure 5: Cross-sectional pairwise correlations among plasma and PiB PET measures at the index visit.*** *. p < .1, * p < .05, ** p < .01, *** p < .001. Aβ, amyloid-β; cDVR, cortical distribution volume ratio; GFAP, glial fibrillary acidic protein; NfL, neurofilament light chain; PiB, Pittsburgh compound B; p-tau, phosphorylated tau.*

#### Longitudinal plasma biomarker and PiB cDVR data used in analyses

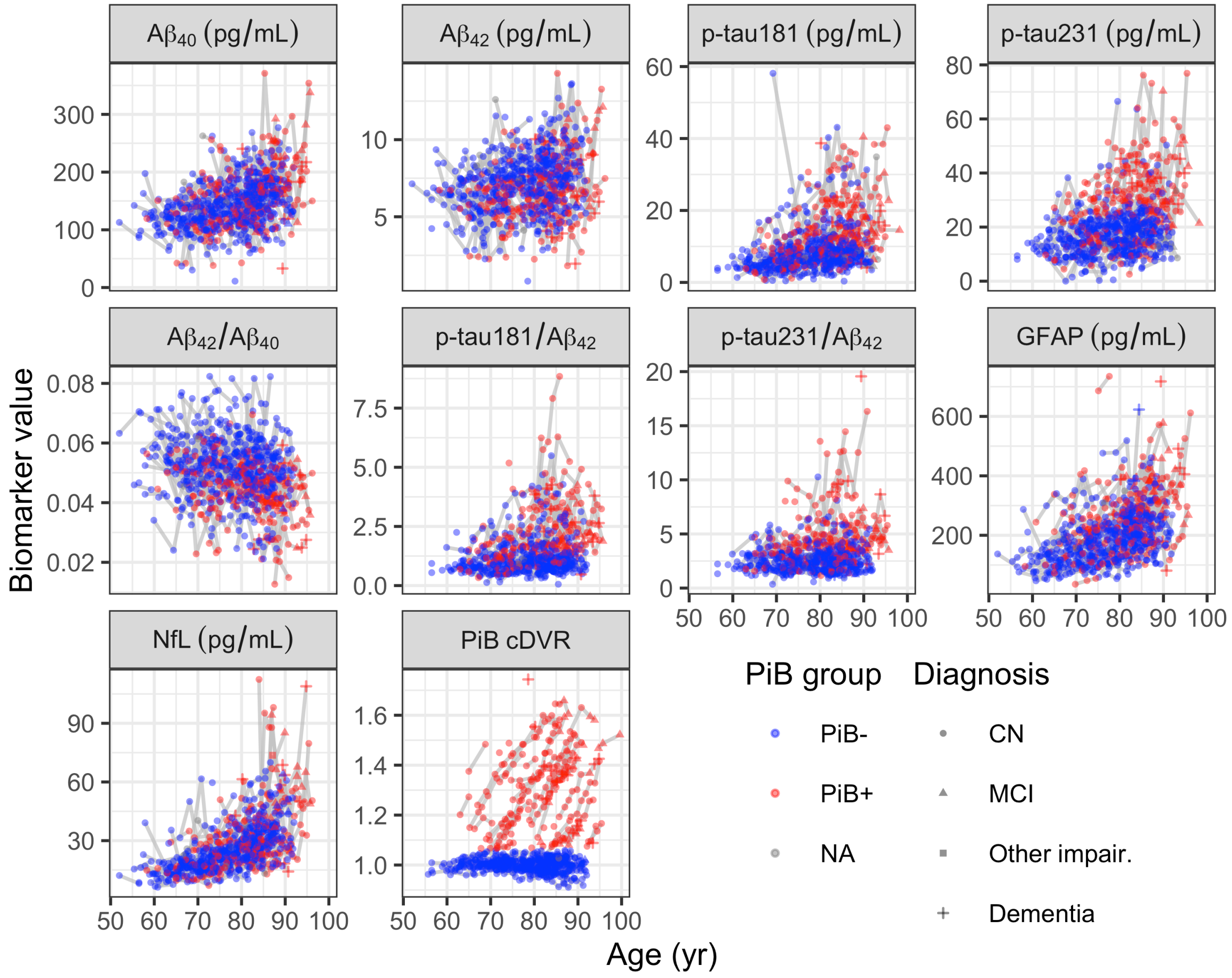


***Supplementary Figure 6: Longitudinal plasma biomarkers and PiB cDVR versus age.*** *Aβ, amyloid-β; cDVR, cortical distribution volume ratio; GFAP, glial fibrillary acidic protein; NfL, neurofilament light chain; PiB, Pittsburgh compound B; p-tau, phosphorylated tau.*

#### Classification of brain amyloid status using plasma biomarkers

***Supplementary Table 1: AUCs for univariate models for predicting PiB group.***

| Variable | AUC | 95% CI |
|---|---|---|
| Age | 0.63 | (0.54–0.71) |
| A$\beta_{40}$ | 0.62 | (0.53–0.7) |
| A$\beta_{42}$ | 0.56 | (0.47–0.65) |
| p-tau181 | 0.72 | (0.63–0.8) |
| p-tau231 | 0.76 | (0.67–0.85) |
| A$\beta_{42}$/A$\beta_{40}$ | 0.72 | (0.65–0.79) |
| p-tau181/A$\beta_{42}$ | 0.77 | (0.7–0.84) |
| p-tau231/A$\beta_{42}$ | 0.78 | (0.71–0.86) |
| GFAP | 0.71 | (0.63–0.79) |
| NfL | 0.64 | (0.55–0.72) |

Abbreviations: Aβ, amyloid-beta; AUC, area under the receiver operating characteristic curve; CI, confidence interval; GFAP, glial fibrillary acidic protein; NfL, neurofilament light chain; p-tau, phosphorylated tau.

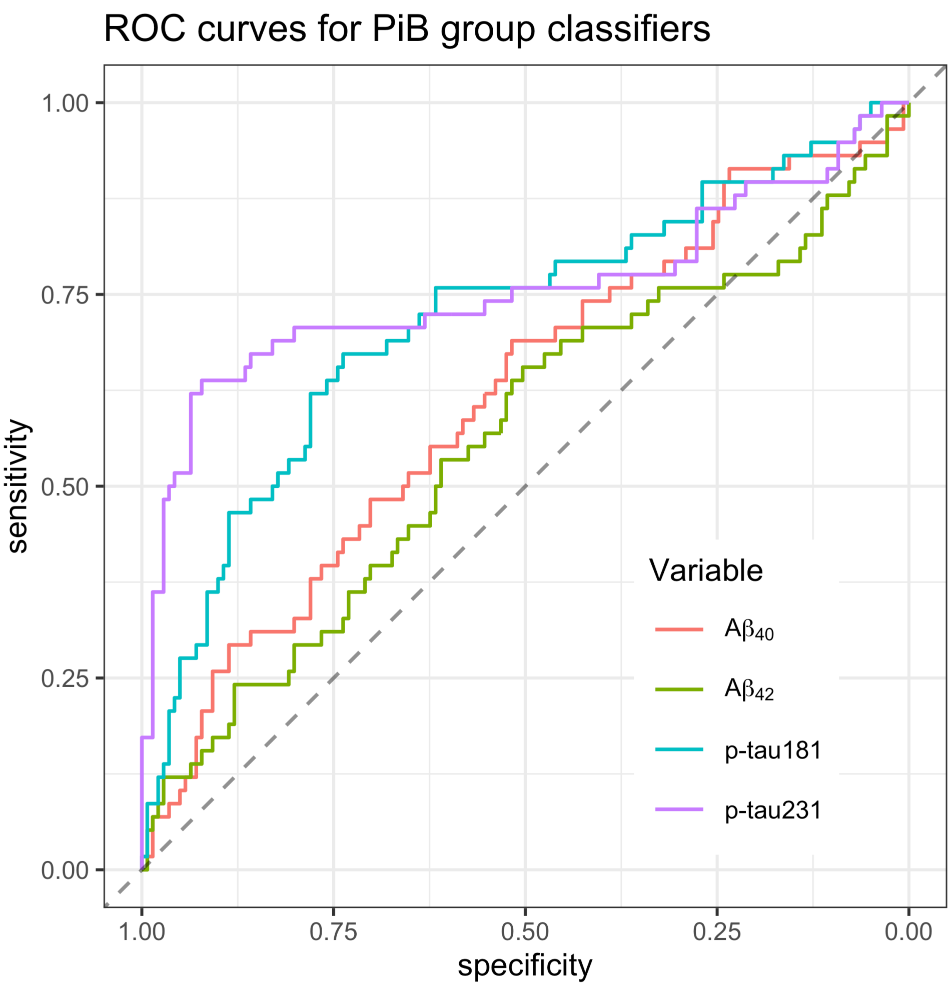


***Supplementary Figure 7: Receiver operating characteristic curves for univariate models for predicting PiB group.*** *Aβ, amyloid-β; PiB, Pittsburgh compound B; p-tau, phosphorylated tau; ROC, receiver operating characteristic.*

#### Longitudinal intraclass correlation coefficients

***Supplementary Table 2: Longitudinal intraclass correlation coefficients.***

| **Biomarker** | **Overall** | | **PiB−** | | **PiB+** | |
|---|---|---|---|---|---|---|
| | **ICC** | **95% CI** | **ICC** | **95% CI** | **ICC** | **95% CI** |
| $A\beta_{40}$ | 0.48 | (0.39–0.55) | 0.47 | (0.35–0.58) | 0.64 | (0.5–0.75) |
| $A\beta_{42}$ | 0.49 | (0.41–0.57) | 0.59 | (0.48–0.68) | 0.76 | (0.64–0.84) |
| p-tau181 | 0.62 | (0.55–0.68) | 0.61 | (0.51–0.7) | 0.80 | (0.7–0.87) |
| p-tau231 | 0.68 | (0.61–0.74) | 0.61 | (0.51–0.7) | 0.80 | (0.7–0.87) |

Abbreviations: Aβ, amyloid-beta; cDVR, cortical distribution volume ratio; CI, confidence interval; GFAP, glial fibrillary acidic protein; ICC, intraclass correlation coefficient; NfL, neurofilament light chain; PiB, Pittsburgh compound B; p-tau, phosphorylated tau.

#### Longitudinal plasma biomarker trajectories by brain amyloid status

We calculated standardized coefficients for the associations reported in Supplementary Tables 3a and 4a by dividing the estimates and their standard errors by the standard deviation of the corresponding outcome measure at the index visit. This standardization yields regression coefficients in units of plasma biomarker *z*-score for level at index visit (Supplementary Table 3b) or plasma biomarker *z*-score per year for longitudinal rates (Supplementary Table 4b).

The data provided in Supplementary Tables 3 and 4 allow for the calculation of annual percent change based on the LMEM results. For example, the percent annual relative change in $A\beta_{42}/A\beta_{40}$ among PiB– can be calculated by dividing -0.000385 (the slope among PiB– from the adjusted model, Supplementary Table 4a) by 0.0527 (the intercept among PiB– from the adjusted model, Supplementary Table 3a), which is -0.73%.

***Supplementary Table 3a: Linear mixed effects model results for intercept.*** *A linear mixed effects model was fitted per biomarker. Models included PiB group at index visit, time from index visit, and their interaction, allowing for the calculation of an average biomarker trajectory per PiB group. Adjusted models additionally covaried for age at index visit, sex, race, APOE ε4 status, and age × time interaction. Post-hoc estimates of biomarker levels at index visit by PiB group are reported. Standard errors are shown in parentheses. * p < .05, ** p < .01, *** p < .001.*

| Biomarker | Model | PiB- | PiB+ | Difference |
|---|---|---|---|---|
| $A\beta_{40}$ | Unadjusted | 138 (2.81)*** | 153 (4.46)*** | 15.5 (5.27)** |
| | Adjusted | 143 (2.31)*** | 148 (3.71)*** | 5.29 (4.41) |
| $A\beta_{42}$ | Unadjusted | 7.11 (0.125)*** | 6.63 (0.197)*** | -0.485 (0.233)* |
| | Adjusted | 7.27 (0.119)*** | 6.45 (0.19)*** | -0.813 (0.227)*** |
| p-tau181 | Unadjusted | 8.21 (0.536)*** | 12.9 (0.835)*** | 4.68 (0.992)*** |
| | Adjusted | 8.75 (0.472)*** | 11.9 (0.75)*** | 3.17 (0.896)*** |
| p-tau231 | Unadjusted | 16.1 (0.705)*** | 27.1 (1.1)*** | 11.1 (1.31)*** |
| | Adjusted | 16.8 (0.652)*** | 26.2 (1.04)*** | 9.41 (1.24)*** |
| $A\beta_{42}/A\beta_{40}$ | Unadjusted | 0.0531 (0.000752)*** | 0.0445 (0.00118)*** | -0.0086 (0.0014)*** |
| | Adjusted | 0.0527 (0.000741)*** | 0.0451 (0.00118)*** | -0.00758 (0.00141)*** |
| p-tau181/$A\beta_{42}$ | Unadjusted | 1.21 (0.074)*** | 1.99 (0.115)*** | 0.78 (0.137)*** |
| | Adjusted | 1.26 (0.0681)*** | 1.86 (0.107)*** | 0.599 (0.129)*** |
| p-tau231/$A\beta_{42}$ | Unadjusted | 2.47 (0.129)*** | 4.46 (0.2)*** | 1.99 (0.238)*** |
| | Adjusted | 2.51 (0.128)*** | 4.36 (0.202)*** | 1.86 (0.243)*** |
| GFAP | Unadjusted | 178 (6.78)*** | 247 (10.6)*** | 68.9 (12.6)*** |
| | Adjusted | 186 (6.08)*** | 230 (9.63)*** | 44.1 (11.6)*** |
| NfL | Unadjusted | 23.6 (0.919)*** | 29.4 (1.44)*** | 5.87 (1.71)*** |
| | Adjusted | 25.1 (0.711)*** | 26.8 (1.13)*** | 1.73 (1.35) |

Abbreviations: Aβ, amyloid-beta; GFAP, glial fibrillary acidic protein; NfL, neurofilament light chain; PiB, Pittsburgh compound B; p-tau, phosphorylated tau.

***Supplementary Table 3b: Linear mixed effects model results (standardized coefficients) for intercept.***

*This table shows the same results as Supplementary Table 3a, except that the estimates have been divided by the standard deviation of the outcome plasma measure at index visits to yield standardized coefficients. Standard errors are shown in parentheses. * p < .05, ** p < .01, *** p < .001.*

| Biomarker | Model | PiB- | PiB+ | Difference |
|---|---|---|---|---|
| $A\beta_{40}$ | Unadjusted | -0.103 (0.0656) | 0.258 (0.104)* | 0.361 (0.123)** |
| | Adjusted | 0.0116 (0.0538) | 0.135 (0.0864) | 0.123 (0.103) |
| $A\beta_{42}$ | Unadjusted | 0.113 (0.0644) p = 0.08 | -0.137 (0.102) | -0.251 (0.12)* |
| | Adjusted | 0.193 (0.0617)** | -0.227 (0.0983)* | -0.42 (0.118)*** |
| p-tau181 | Unadjusted | -0.193 (0.0772)* | 0.481 (0.12)*** | 0.674 (0.143)*** |
| | Adjusted | -0.115 (0.068) p = 0.09 | 0.341 (0.108)** | 0.456 (0.129)*** |
| p-tau231 | Unadjusted | -0.376 (0.0677)*** | 0.686 (0.106)*** | 1.06 (0.125)*** |
| | Adjusted | -0.31 (0.0626)*** | 0.593 (0.0997)*** | 0.903 (0.119)*** |
| $A\beta_{42}/A\beta_{40}$ | Unadjusted | 0.284 (0.0691)*** | -0.507 (0.109)*** | -0.791 (0.129)*** |
| | Adjusted | 0.242 (0.0681)*** | -0.455 (0.108)*** | -0.697 (0.13)*** |
| p-tau181/$A\beta_{42}$ | Unadjusted | -0.22 (0.0775)** | 0.597 (0.12)*** | 0.817 (0.143)*** |
| | Adjusted | -0.163 (0.0713)* | 0.464 (0.112)*** | 0.627 (0.135)*** |
| p-tau231/$A\beta_{42}$ | Unadjusted | -0.319 (0.067)*** | 0.714 (0.104)*** | 1.03 (0.124)*** |
| | Adjusted | -0.299 (0.0667)*** | 0.666 (0.105)*** | 0.965 (0.127)*** |
| GFAP | Unadjusted | -0.221 (0.0734)** | 0.525 (0.115)*** | 0.746 (0.136)*** |
| | Adjusted | -0.133 (0.0659)* | 0.345 (0.104)** | 0.478 (0.125)*** |
| NfL | Unadjusted | -0.145 (0.0706)* | 0.305 (0.111)** | 0.451 (0.131)*** |
| | Adjusted | -0.0296 (0.0546) | 0.104 (0.0869) | 0.133 (0.104) |

Abbreviations: Aβ, amyloid-beta; GFAP, glial fibrillary acidic protein; NfL, neurofilament light chain; PiB, Pittsburgh compound B; p-tau, phosphorylated tau.

***Supplementary Table 4a: Linear mixed effects model results for longitudinal rates of change in plasma measures by PiB group.*** *A linear mixed effects model was fitted per biomarker. Models included PiB group at index visit, time from index visit, and their interaction, allowing for the calculation of an average biomarker trajectory per PiB group. Adjusted models additionally covaried for age at index visit, sex, race, APOE ε4 status, and age × time interaction. Post-hoc estimates of longitudinal rate of change in biomarkers by PiB group are reported. Standard errors are shown in parentheses. * p < .05, ** p < .01, *** p < .001. p-values between .05 and .1 are stated below. p-values greater than .1 are not indicated.*

| Biomarker | Model | PiB- | PiB+ | Difference |
|---|---|---|---|---|
| $A\beta_{40}$ | Unadjusted | 0.362 (0.602) | 2.36 (1.03)* | 2 (1.2) p = 0.1 |
| | Adjusted | 0.656 (0.557) | 1.17 (0.966) | 0.511 (1.12) |
| $A\beta_{42}$ | Unadjusted | -0.0496 (0.0257) p = 0.06 | 0.11 (0.0439)* | 0.16 (0.0508)** |
| | Adjusted | -0.0409 (0.0253) | 0.0643 (0.0432) | 0.105 (0.0504)* |
| p-tau181 | Unadjusted | 0.0433 (0.0949) | 0.502 (0.152)** | 0.458 (0.18)* |
| | Adjusted | 0.0791 (0.0943) | 0.426 (0.157)** | 0.347 (0.183) p = 0.06 |
| p-tau231 | Unadjusted | 0.339 (0.146)* | 0.957 (0.236)*** | 0.618 (0.277)* |
| | Adjusted | 0.382 (0.143)** | 0.756 (0.24)** | 0.373 (0.278) |
| $A\beta_{42}/A\beta_{40}$ | Unadjusted | -0.000406 (0.0000946)*** | 0.000138 (0.000165) | 0.000544 (0.00019)** |
| | Adjusted | -0.000385 (0.0000977)*** | 0.000156 (0.000167) | 0.000541 (0.000195)** |
| p-tau181/$A\beta_{42}$ | Unadjusted | 0.00397 (0.017) | 0.0602 (0.0273)* | 0.0562 (0.0321) p = 0.08 |
| | Adjusted | -0.000981 (0.0174) | 0.0653 (0.028)* | 0.0663 (0.0336) p = 0.05 |
| p-tau231/$A\beta_{42}$ | Unadjusted | 0.0251 (0.0261) | 0.112 (0.0425)** | 0.0871 (0.0499) p = 0.08 |
| | Adjusted | 0.0229 (0.0268) | 0.11 (0.0439)* | 0.0874 (0.0525) p = 0.1 |
| GFAP | Unadjusted | 4.92 (1.13)*** | 9.16 (1.89)*** | 4.24 (2.21) p = 0.06 |
| | Adjusted | 5.51 (1.13)*** | 7.83 (1.89)*** | 2.32 (2.23) |
| NfL | Unadjusted | 0.838 (0.173)*** | 1.54 (0.292)*** | 0.705 (0.34)* |
| | Adjusted | 0.917 (0.165)*** | 1.14 (0.279)*** | 0.22 (0.327) |

Abbreviations: Aβ, amyloid-beta; GFAP, glial fibrillary acidic protein; NfL, neurofilament light chain; PiB, Pittsburgh compound B; p-tau, phosphorylated tau.

***Supplementary Table 4b: Linear mixed effects model results (standardized coefficients) for longitudinal rates of change in plasma measures by PiB group.*** *This table shows the same results as Supplementary Table 4a, except that the estimates have been divided by the standard deviation of the outcome plasma measure at index visits to yield standardized coefficients. Standard errors are shown in parentheses. * p < .05, ** p < .01, *** p < .001. p-values between .05 and .1 are stated below. p-values greater than .1 are not indicated.*

| Biomarker | Model | PiB- | PiB+ | Difference |
|---|---|---|---|---|
| $A\beta_{40}$ | Unadjusted | 0.00845 (0.014) | 0.0551 (0.0241)* | 0.0466 (0.0279) p = 0.1 |
| | Adjusted | 0.0153 (0.013) | 0.0272 (0.0225) | 0.0119 (0.0262) |
| $A\beta_{42}$ | Unadjusted | -0.0256 (0.0133) p = 0.06 | 0.057 (0.0227)* | 0.0826 (0.0263)** |
| | Adjusted | -0.0211 (0.0131) | 0.0332 (0.0223) | 0.0544 (0.0261)* |
| p-tau181 | Unadjusted | 0.00625 (0.0137) | 0.0723 (0.022)** | 0.0661 (0.0259)* |
| | Adjusted | 0.0114 (0.0136) | 0.0614 (0.0227)** | 0.05 (0.0263) p = 0.06 |
| p-tau231 | Unadjusted | 0.0326 (0.014)* | 0.0919 (0.0226)*** | 0.0593 (0.0266)* |
| | Adjusted | 0.0367 (0.0138)** | 0.0726 (0.0231)** | 0.0359 (0.0267) |
| $A\beta_{42}/A\beta_{40}$ | Unadjusted | -0.0373 (0.0087)*** | 0.0127 (0.0151) | 0.05 (0.0175)** |
| | Adjusted | -0.0354 (0.00898)*** | 0.0144 (0.0153) | 0.0498 (0.018)** |
| p-tau181/$A\beta_{42}$ | Unadjusted | 0.00416 (0.0178) | 0.063 (0.0286)* | 0.0588 (0.0336) p = 0.08 |
| | Adjusted | -0.00103 (0.0182) | 0.0684 (0.0293)* | 0.0694 (0.0351) p = 0.05 |
| p-tau231/$A\beta_{42}$ | Unadjusted | 0.0131 (0.0136) | 0.0583 (0.0221)** | 0.0453 (0.0259) p = 0.08 |
| | Adjusted | 0.0119 (0.014) | 0.0574 (0.0228)* | 0.0455 (0.0273) p = 0.1 |
| GFAP | Unadjusted | 0.0533 (0.0123)*** | 0.0992 (0.0205)*** | 0.0459 (0.0239) p = 0.06 |
| | Adjusted | 0.0597 (0.0123)*** | 0.0847 (0.0205)*** | 0.0251 (0.0241) |
| NfL | Unadjusted | 0.0644 (0.0133)*** | 0.119 (0.0224)*** | 0.0541 (0.0261)* |
| | Adjusted | 0.0704 (0.0127)*** | 0.0873 (0.0215)*** | 0.0169 (0.0251) |

Abbreviations: Aβ, amyloid-beta; GFAP, glial fibrillary acidic protein; NfL, neurofilament light chain; PiB, Pittsburgh compound B; p-tau, phosphorylated tau.

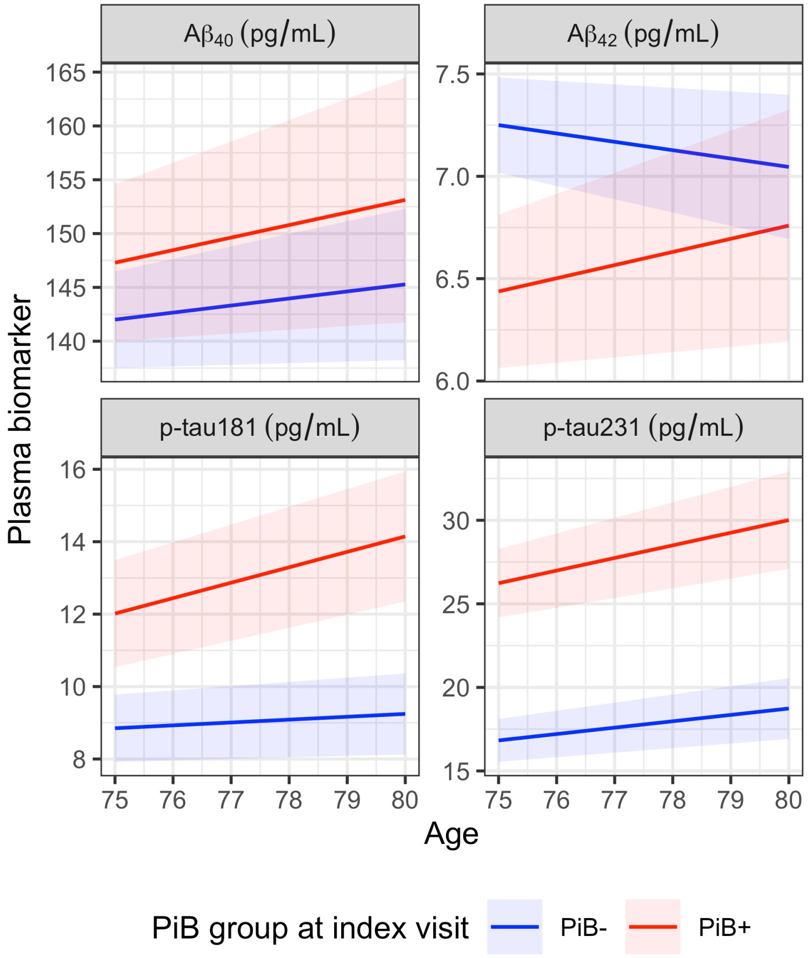


***Supplementary Figure 8: Predicted plasma biomarker trajectories for A$\beta_{40}$, A$\beta_{42}$, p-tau181, and p-tau231.*** *A$\beta$, amyloid-$\beta$; PiB, Pittsburgh compound B; p-tau, phosphorylated tau.*

#### Associations among longitudinal rates of change in plasma biomarkers and brain amyloid

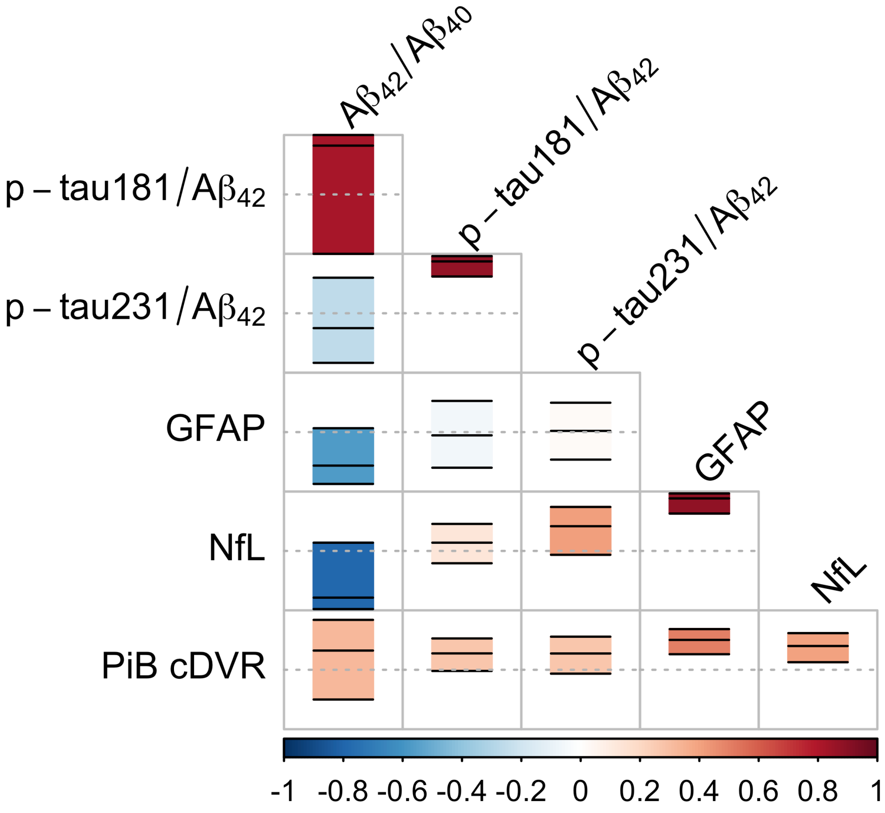


***Supplementary Figure 9: Pairwise correlations among rates of change of plasma and PiB PET measures, as assessed using the random effect correlation matrix estimated in the bivariate linear mixed effects models.*** *The rectangle in each cell indicates the 95% confidence interval of the correlation estimate, which is shown with a black solid line inside the rectangle. Dashed horizontal lines correspond to a correlation of 0. Color indicates the correlation estimate. Aβ, amyloid-β; cDVR, cortical distribution volume ratio; GFAP, glial fibrillary acidic protein; NfL, neurofilament light chain; PiB, Pittsburgh compound B; p-tau, phosphorylated tau.*

#### Temporal order of changes in plasma biomarkers and brain amyloid

##### Progression score model

The progression score (PS) model used in our analysis is given by

$$
\begin{aligned}
s_{ij} &= t_{ij} + \tau_i \\
y_{ijk} &= a_k \,\mathrm{logit}^{-1}\big(b_k(s_{ij} - c_k)\big) + d_k + \epsilon_{ijk} \\
\tau_i &\sim \mathcal{N}(0, \sigma_\tau^2) \\
\boldsymbol{\epsilon}_{ij} &\sim \mathcal{N}(0, \boldsymbol{\Sigma}),
\end{aligned}
$$

where $t_{ij}$ is age (or time) of subject $i$ at visit $j$, $\tau_i$ is the time-shift for this subject, $s_{ij}$ is the PS at this visit, $y_{ijk}$ is the measurement for $k^{\text{th}}$ biomarker at this visit, $\{a_k, b_k, c_k, d_k\}$ are sigmoid trajectory parameters for this biomarker, and $\epsilon_{ijk}$ is random noise. The modeling of correlations among biomarkers through the covariance matrix $\boldsymbol{\Sigma}$ ensures that PS will not be unduly influenced by overlapping information conveyed by the biomarkers.

For simplicity in model fitting, we assumed that all biomarker trajectories were increasing, which allowed us to use more specific hyperpriors for the sigmoid to guide parameter estimation. (Given this assumption, $\mathrm{A}\beta_{42}/\mathrm{A}\beta_{40}$ values were negated prior to model fitting, and results were transformed back to the original scale of the biomarker.)

Selection of hyperpriors were guided in part by [2] and the Bayesian linear regression implementation in `rstanarm`. Hyperpriors for the sigmoid parameters were

$$
\begin{aligned}
a_k &\sim \text{Half-}\mathcal{N}(3.92\,\sigma_{y_k}, \sigma_{a_k}^2) \\
b_k &\sim \text{Half-}\mathcal{N}(0, \sigma_{b_k}^2) \\
c_k &\sim \mathcal{N}(0, (3\sigma_t)^2) \\
d_k &\sim \mathcal{N}(\mu_{y_k} - 1.96\,\sigma_{y_k}, \sigma_{a_k}^2),
\end{aligned}
$$

where $\mu_{y_k}$ and $\sigma_{y_k}$ are the mean and standard deviation of the $k^{\text{th}}$ biomarker, respectively, $\sigma_t$ is the standard deviation of the time variable $t$ (across all subjects and visits), $\sigma_{a_k} = 1.5\,\sigma_{y_k}$, and $\sigma_{b_k} = 1/\sigma_t$.

The biomarker residual covariance matrix was parameterized using $\Sigma = \Lambda C \Lambda$, where $\Lambda$ is a diagonal matrix with positive diagonal entries and $C$ is a positive-definite correlation matrix with the following priors:

$$
\begin{aligned}
[\Lambda]_{kk} &\sim \text{Exp}(\sigma_{y_k}^{-1}) \\
C &\sim \text{LKJ}(1).
\end{aligned}
$$

For the standard deviation of the subject-specific time-shift, we used the hyperprior $\sigma_\tau \sim$ Half-Cauchy$(0{,}5)$.

We estimated model parameters using Hamiltonian Monte Carlo Markov chain with 4 chains and 10,000 warm-up samples followed by 5000 posterior draws per chain using RStan [3]. There were no divergent transitions after warm-up. Maximum $\hat{R}$ was 1.02, indicating model convergence.

#### Time interval between peak relative change in plasma biomarkers and amyloid positivity onset

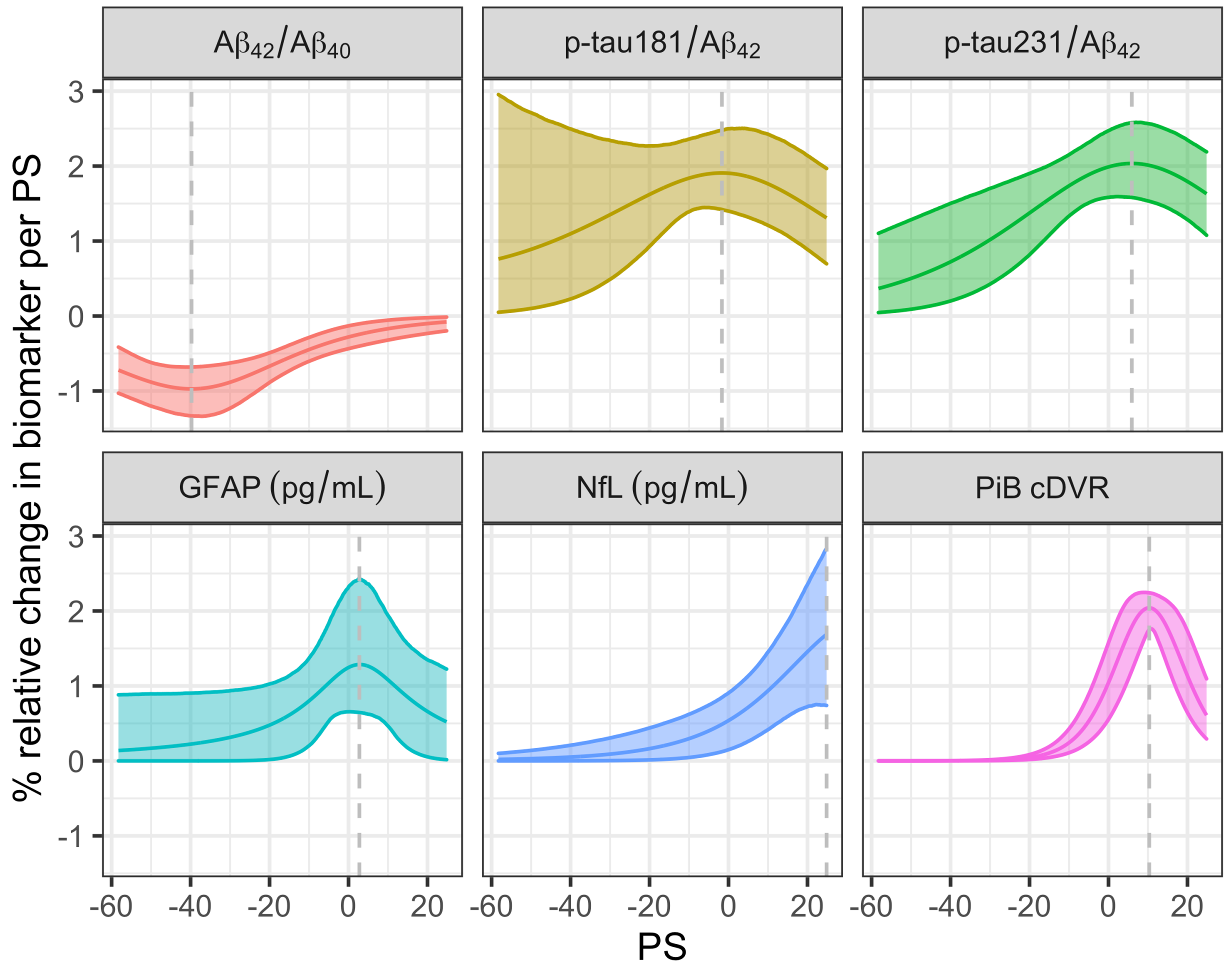


***Supplementary Figure 10: Percent relative change in biomarkers per PS as a function of PS.*** *Bands indicate 95% confidence intervals. Vertical dashed lines indicate the PS at which absolute percent relative change is maximal.*

***Supplementary Table 5: Peak percent relative biomarker change (per year) and time interval (in years) between peak relative change and the PS value that corresponds to PiB positivity threshold.***

| Biomarker | Peak % relative change | Interval between peak and amyloid onset |
|---|---|---|
| $A\beta_{42}/A\beta_{40}$ | -0.99 (-1.37, -0.69) | -41.3 (-53, -31.7) |
| p-tau181/$A\beta_{42}$ | 1.96 (1.56, 3.45) | -5.1 (-37.6, 12.1) |
| p-tau231/$A\beta_{42}$ | 2.08 (1.63, 2.64) | 4.7 (-10.9, 15.5) |
| GFAP | 1.42 (0.74, 2.68) | 1.5 (-26, 18) |
| NfL | 1.98 (0.88, 3.46) | 17.9 (-11.8, 27.1) |
| PiB cDVR | 2.11 (1.95, 2.28) | 10.1 (9.1, 11.2) |

Abbreviations: Aβ, amyloid-beta; cDVR, cortical distribution volume ratio; GFAP, glial fibrillary acidic protein; NfL, neurofilament light chain; PiB, Pittsburgh compound B; p-tau, phosphorylated tau.

#### PS model result using p-tau concentrations rather than p-tau/$A\beta_{42}$

We repeated the PS model using p-tau concentrations rather than p-tau/$A\beta_{42}$. Results are presented in Supplementary Figures 11–13.

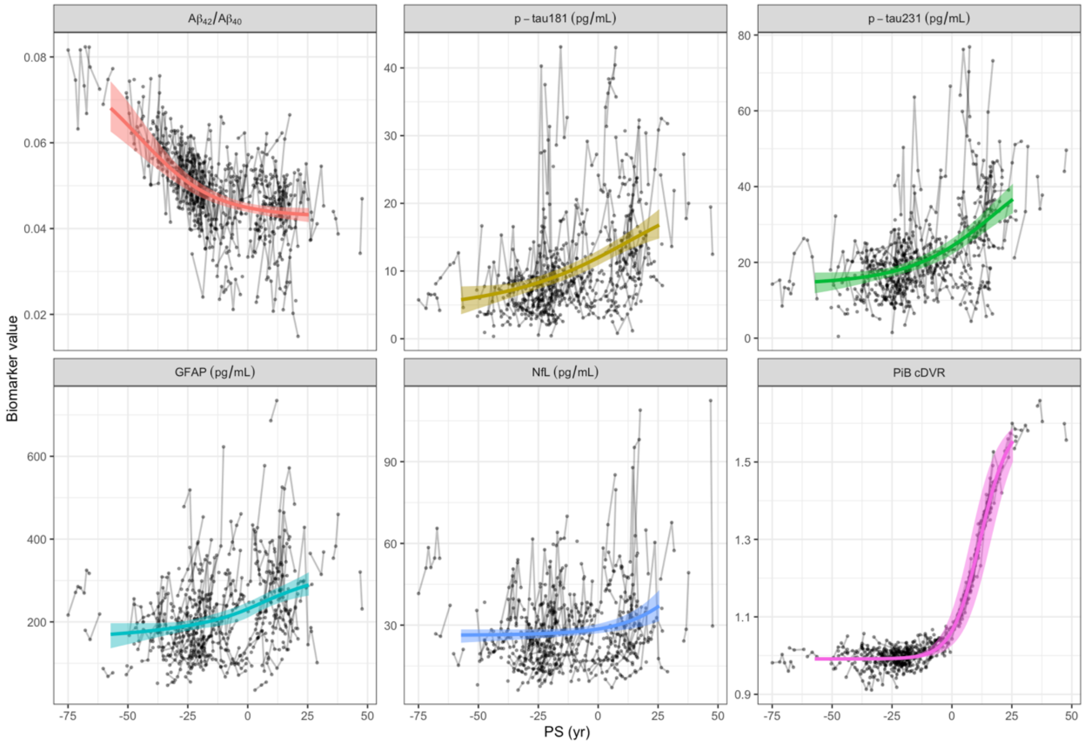


***Supplementary Figure 11: Result of the progression score (PS) model with p-tau concentrations instead of p-tau/$A\beta_{42}$: Biomarker trajectories estimated after alignment of individual-level longitudinal data using the PS model.*** *Bands indicate the 95% confidence intervals for the trajectory estimates. PS scale was calibrated after model fitting such that at PS = 0, the estimated trajectory for PiB cDVR attains the value 1.06, which is the PiB positivity threshold. Since PS is time-shifted age, it is in the units of years. Aβ, amyloid-β; cDVR, cortical distribution volume ratio; GFAP, glial fibrillary acidic protein; NfL, neurofilament light chain; PiB, Pittsburgh compound B; PS, progression score; p-tau, phosphorylated tau.*

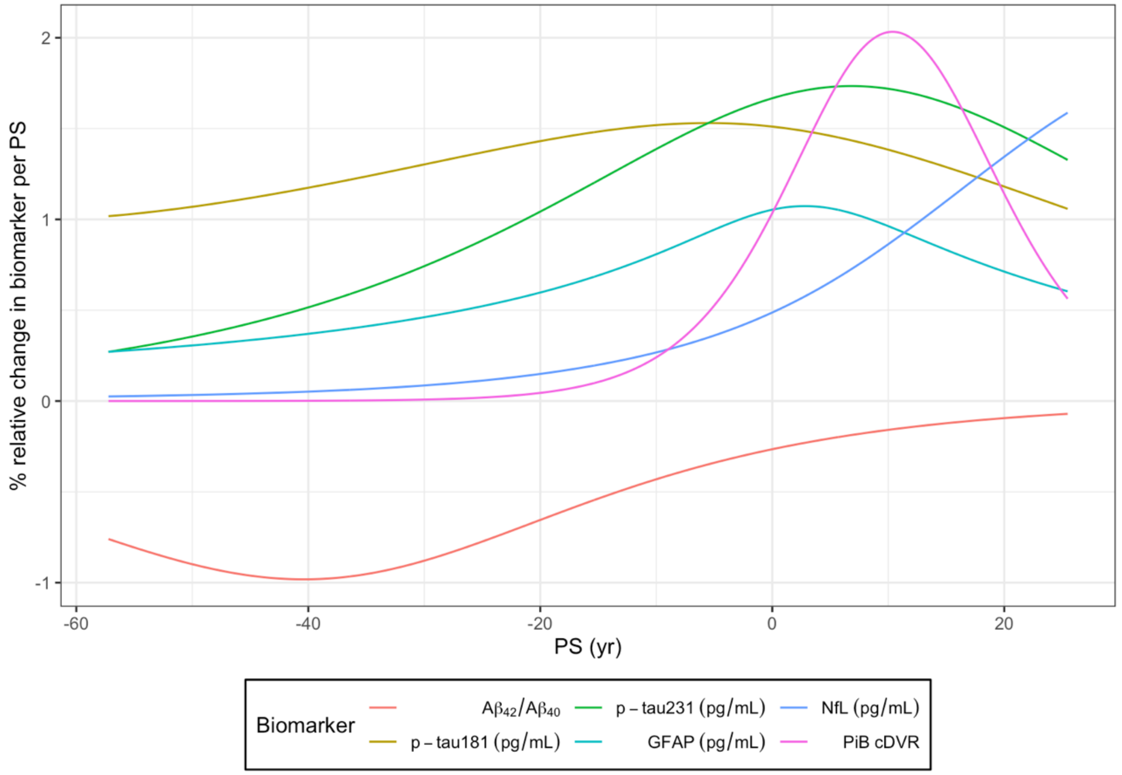


***Supplementary Figure 12: Result of the progression score (PS) model with p-tau concentrations instead of p-tau/Aβ$_{42}$: Percent relative change in biomarkers per PS as a function of PS.*** *Aβ, amyloid-β; cDVR, cortical distribution volume ratio; GFAP, glial fibrillary acidic protein; NfL, neurofilament light chain; PiB, Pittsburgh compound B; PS, progression score; p-tau, phosphorylated tau.*

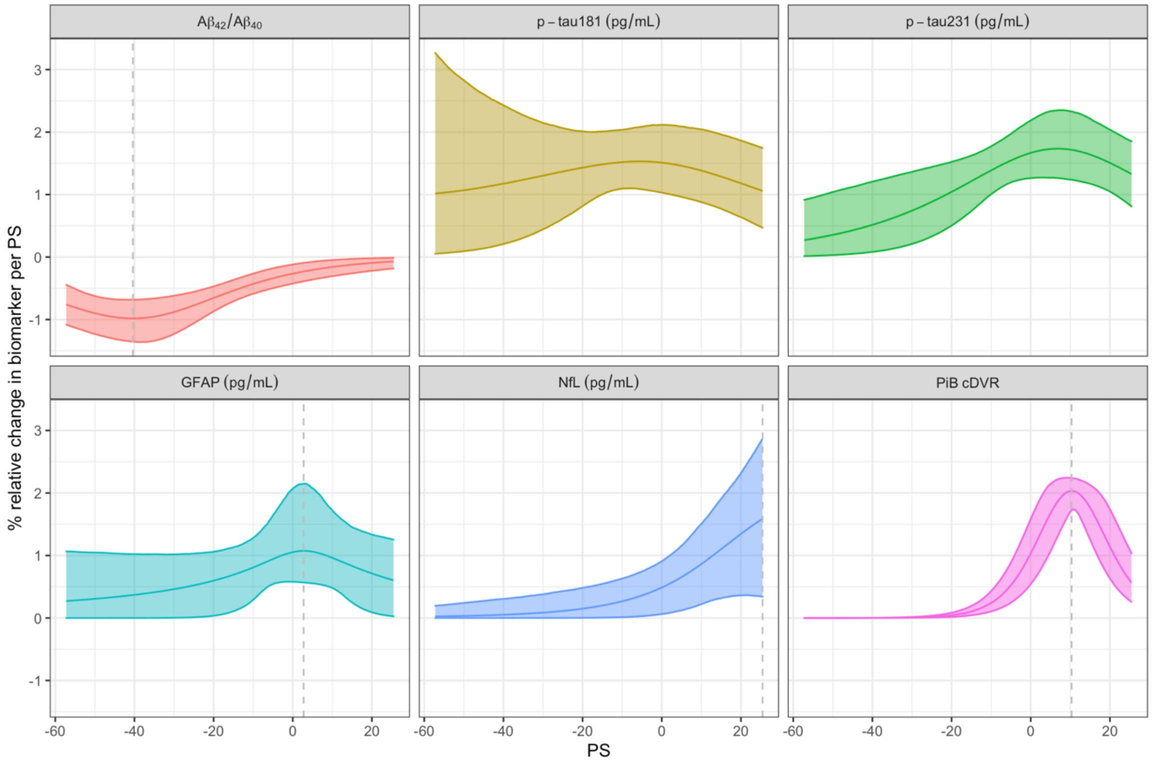


***Supplementary Figure 13: Result of the progression score (PS) model with p-tau concentrations instead of p-tau/A$\beta_{42}$: Percent relative change in biomarkers per PS as a function of PS.*** *Bands indicate 95% confidence intervals. Vertical dashed lines indicate the PS at which absolute percent relative change is maximal.*
